# Evaluating Open-Source Wrist-Worn Accelerometer Models for Sedentary Time Detection Against Thigh-Worn Accelerometer Data

**DOI:** 10.64898/2026.06.30.26356834

**Authors:** Aidan Acquah, Katya Broomberg, David W Dunstan, Genevieve N. Healy, Melanie J. Davies, Charlotte L. Edwardson, Aiden Doherty, Benjamin D. Maylor

## Abstract

**Objective:** Wrist-worn accelerometers are common in large-scale epidemiological studies, but their ability to measure sedentary behaviour in free-living environments is unknown. We therefore aimed to evaluate the accuracy of openly-available methods to infer sedentary time from wrist-worn accelerometers.

**Methods:** We analysed data from 662 working-age adults in the SMART Work & Life study (20-70 years; mean age 45 years; 72% female) who concurrently wore wrist- and thigh-worn accelerometers for up to eight free-living days. Reference measurements of sedentary time were derived from the thigh accelerometer data using proprietary algorithms. Wrist accelerometer data were processed using widely used, publicly available activity recognition models. Performance was evaluated at 30-second epochs to generate per-participant metrics, alongside comparisons of mean daily sedentary time, mean daily number of prolonged sedentary bouts (≥ 30 minutes) and proportion of sedentary time in prolonged bouts. Model performance was examined across subgroups defined by age, sex, body mass index, season, recruitment centre, and in sensitivity analyses restricted to daytime hours (08:00–22:00).

**Results:** The best performing machine learning model (Actinet) accurately classified sedentary time from wrist-worn accelerometer data with a mean per-participant accuracy of 0.87 and F1 score of 0.85. Cut point-based approaches demonstrated lower accuracy of 0.80 (F1 score of 0.79). The ActiNet machine learning model showed strong agreement in daily sedentary time, daily number of prolonged sedentary bouts and proportion of sedentary time in prolonged bouts, all within 10% of the free-living thigh reference. Findings were consistent across subgroups and in analyses restricted to daytime hours.

**Conclusion:** Wrist-worn accelerometers can provide accurate measurements of sedentary behaviour in free-living settings, when assessed using current machine learning models, particularly ActiNet. This work provides confidence in future epidemiological research to examine sedentary behaviour patterns from wrist-worn accelerometers and their associations with health outcomes.

## Introduction

High amounts of time spent sedentary, defined as waking behaviour in a seated, reclined, or lying position,^1^ is associated with a range of adverse health outcomes, including cardiometabolic disease, type 2 diabetes, and premature mortality. ^2–8^ Beyond the average duration of daily sedentary time, emerging evidence suggests that more detailed characteristics of sedentary behaviour may also be important for health. These include differences between weekdays and weekends, ^9,10^ between daytime and night-time periods,^11,12^ and patterns of accumulation, such as the time accrued, and the average duration of sedentary bouts^13–15^. The increasing use of wearable monitoring devices has enabled objective measurement of movement behaviours, reducing the recall error and social desirability bias inherent to self-reported measures^16,17^. Thigh-worn accelerometers can be used as a “silver standard” in lieu of direct observation for sedentary behaviour detection and can distinguish sitting or lying from upright postures^18–21^. In contrast, wrist-worn accelerometers typically infer sedentary behaviour from low movement intensity rather than posture^22^. Despite this limitation, wrist-worn devices are more economical and widely adopted in both large-scale epidemiological studies and consumer settings such as smartwatches, ^23,24^, making them far more scalable for population health research.

However, it remains unclear how wrist-worn accelerometer activity recognition models perform relative to a thigh-worn accelerometer reference in large free-living populations.^22,25^ Although studies such as Matthews et al. (2025)^26^ and Pavey et al. (2016)^27^ have demonstrated that mean daily sedentary time estimates derived from wrist-worn models are broadly equivalent to those obtained from thigh-worn monitors, these findings are drawn from small samples and may not generalise more widely. Beyond daily-level agreement, epoch-level construct validity and the equivalency of derived sedentary bout metrics have received comparatively little attention. Finally, model failure cases remain poorly characterised, including differential performance across subgroups and the misclassification of behaviours such as sleep, where low-amplitude wrist signals can closely resemble sedentary activity^25^.

In this study, we therefore evaluated the construct validity of sedentary behaviour detected from wrist-worn accelerometers using several open-source activity recognition models, with thigh-worn accelerometer data serving as the reference standard. We assessed performance at multiple levels, including per-participant agreement for aligned 30-second activity epochs, equivalence in average daily sedentary time, average daily number of prolonged sedentary bouts, and average proportion of sedentary behaviour in prolonged sedentary bouts. We additionally examined how these models performed, within subgroups of interests, such as age and sex, as well as considering only waking day hours of 08:00AM to 10:00PM, to avoid the misclassification of sleep. The results of these analyses may inform the choice of wrist-accelerometer model, and derived digital measures selected for the monitoring of sedentary behaviour in future epidemiological analyses.

## Methods

### Study population

These analyses used baseline data from the SMART Work & Life (SWAL) study^28^, a cluster randomised controlled trial of an intervention designed to reduce sitting in desk-based employees conducted in England, between May 2018 and February 2019. All participants provided informed consent prior to participation, and the study received appropriate ethical approval from two research sites (Leicester ref:14372; Salford ref:HSR1718-039).

A total of 756 participants were enrolled in the study at baseline, during which participants wore two accelerometers simultaneously for 8 days during free-living; a thigh-worn activPAL, and a wrist-worn AX3 Axivity. Devices were worn continuously (24 hours a day) according to established study protocols. Demographic data, including age and sex, were self-reported by participants from questionnaires, while body mass index (BMI) was obtained through measured height and fasting body mass^28^.

Participants were excluded from analyses if: (1) Non-unique IDs were found in the raw wrist or thigh dataset (likely due to a device failure at the first attempt, leading to re-monitoring), (2) Corresponding IDs were not found between raw wrist and thigh datasets, (3) Models failed to extract valid outputs (e.g., due to calibration failure or algorithm malfunction), (4) Demographic data was missing, or (5) Timestamps from the thigh and wrist monitors did not align (i.e., zero minutes of concurrent data).

### Activity Recognition Models

#### Thigh

The thigh monitor used in this study was an activPAL3 micro (PAL Technologies Ltd., Glasgow, UK), collecting triaxial accelerometry at 20Hz with a dynamic range of ±2*g*. Raw data were processed using the proprietary PALbatch software (v9.1.3.94; CREA algorithm v1.3)^29^, generating 1-second activity classifications for time spent primary lying (asleep), secondary lying (non-sleep lying), sedentary (non-vehicle sitting), vehicle (vehicle sitting), upright (standing), stepping (walking) and bicycling. For this analysis, these labels were collapsed into a binary classification of sedentary versus non-sedentary behaviour. Sedentary behaviour was defined as epochs classified as secondary lying, sedentary, or vehicle, while all remaining labels (primary lying, upright, stepping, bicycling) were classified as non-sedentary.

This classification algorithm uses device orientation, relative to gravity, to infer posture and has demonstrated strong performance for posture-based behaviours (sitting, standing, walking) in free-living and direct observation studies^22,30^. The PALbatch outputs were treated as the reference standard for sedentary behaviour, consistent with thigh-worn accelerometers’ broader proposed role as a traceable secondary reference measure for field-based sedentary time assessment. ^31^

#### Wrist

The wrist monitor was an Axivity AX3 (Axivity Ltd., Newcastle upon Tyne, UK) worn on the non-dominant wrist, collecting triaxial acceleration at 100Hz with a dynamic range of ±8*g*. The raw data files were processed using existing, open access, activity recognition approaches, selected based on good equivalency of extracted daily sedentary time, reported by Matthews et al. (2025)^26^, alongside one recently released model with reported classification performance improvements^32^.

##### Machine learning models

The Accelerometer model, implemented through the python package ‘accelerometer’ version 7.5.0^33^ applies a balanced random forest classifier followed by hidden Markov model (HMM) smoothing. Data were gravity-calibrated and segmented into non-overlapping 30-second windows. Features extracted from each window were classified into sleep, sedentary, light, or moderate-to-vigorous activity, with HMM smoothing applied to produce the most likely state sequence. A one-hour sleep correction reclassified sleep bouts shorter than one hour as sedentary^34^.

The ActiNet model, implemented through the python package ‘actinet’ version 0.7.0^35^ uses a HARNet model^36^, followed by HMM smoothing. Data were gravity-calibrated, linearly downsampled to 30 Hz, and segmented into 30-second non-overlapping windows. Classification labels matched those of the ‘accelerometer’ model, with identical one-hour sleep correction procedures.^32^

The Actimetric model, available through ‘actimetric’ R package GitHub repository version 0.1.5^37^ consists of a random forest model, producing activity labels to 10-second windows of data, using the GGIR package for data reading and gravity calibration^38,39^. We use the extended version of this algorithm, proposed by Ahmadi et al. (2020), to incorporate sleep detection^40^. The output labels from this extended model were nighttime.sleep, nighttime.awake, sedentary, stationary, walk and run, with binary sedentary labelled for ‘sedentary’ with all other labels mapping on to ‘not-sedentary’.

##### Cutpoint-based models

Cutpoint-based models were implemented through the ‘GGIR’ R package version 3.2-7^39^. Raw accelerometry data were gravity-calibrated and segmented into 5-second epochs. Acceleration magnitude was summarised using either Euclidean Norm Minus One (ENMO) or Euclidean Norm Minus One Absolute (ENMOabs)^41^ depending on the model.

Classification labels of inactive (treated as sedentary in this analysis) and light-intensity activity were distinguished based on exceeding cutpoint values of the acceleration magnitude measure, specific to model. The GGIR default model used an ENMO threshold of 40mg, the Bakrania model used an ENMO threshold of 32.8mg^42^, and the Fraysse model used an ENMOabs threshold of 62.5mg^43^. Sleep detection was performed using heuristic and sustained inactivity detection algorithms^44,45^.

The python-based activity recognition models (Accelerometer and ActiNet) used python version 3.9, while the R-based models (Actimetric and all GGIR models) used R version 4.3.2.

### Model Harmonisation and Alignment

To enable direct comparison, all model outputs were harmonised to 30-second epochs, starting at midnight on the first monitoring day. For models producing higher-resolution outputs (5-second or 10-second epochs), the modal label within each 30-second window was assigned. For PALbatch outputs, behaviour durations were summed within each 30-second window, and the dominant label (>15 seconds) was assigned. If any model produced missing or invalid classifications for a given epoch, that epoch was excluded from all model comparisons for that participant.

### Model evaluation

#### Epoch-Level Agreement

Agreement between outputs from the wrist-based models, and the thigh-reference was evaluated at the epoch level. Metrics included accuracy, binary F1 score, Cohen’s kappa, sensitivity and specificity for sedentary classification, with the average performances provided for all study participants. Confusion matrices, as well as cross tabulations for the binary and full activity labels, respectively, were provided, to identify failure cases of the model.

#### Participant-Level Agreement

We also assessed how the models compared in the agreement of derived sedentary metrics, as compared to the reference standard. We first extracted mean daily sedentary time, a measure of the total amount of detected sedentary time on each calendar day, averaged across all days of monitoring, without any imputation of missing data.

Beyond the measurement of the equivalency of the aggregation of sedentary time, we also extracted and compared the equivalency of sedentary time fragmentation. To do so, we calculated two prolonged sedentary bout related metrics: average daily number of prolonged bouts, and the average proportion of total sedentary time spent in prolonged bouts. In this study, we define a prolonged sedentary bout as distinct periods of detected sedentary behaviour, lasting at least 30 minutes, with a 90-second tolerance for non-sedentary gaps (i.e. a maximum of three consecutive 30-second windows of non-sedentary behaviour), and at least 80% of the distinct period classified as sedentary, informed by existing literature^15,46^.

Agreement was compared using equivalency tests. Differences between wrist and thigh-derived metrics were calculated, and equivalence was observed if the mean difference and 90% confidence interval fell within ±10% of the thigh-derived mean value for that metric. Additionally, digital measures extracted from wrist-based models were compared with thigh-derived values using root mean squared error (RMSE), mean absolute error (MAE), and the Pearson correlation coefficient.

### Subgroup Analyses

We investigated how the performance of models differed within subgroups of participants, that may have different patterns of behaviour. These groups included age (<35, 35-45, 45-55, 55+), sex (male, female), BMI (underweight/normal, overweight, obese), season of device wear (spring, summer, autumn, winter), and recruitment centre (Leicester, Salford).

Within each of these subgroups, we evaluated the epoch-level agreement as outlined above.

### Sensitivity Analysis

Due to known challenges in distinguishing wake from sleep periods using a thigh monitor^47,48^, we conducted a sensitivity analysis, restricting analyses to presumed waking hours of between 08:00 AM and 10:00 PM, a more conservative estimate of the waking day from prior literature (07:00 AM to 11:00 PM)^49^. Within this sensitivity analysis, we evaluated the epoch- and feature-level agreements between the wrist and thigh models, while reducing the reliance on accurate sleep classification.

These analyses were run using python version 3.14, scikit-learn version 1.8.0.

## Results

### Study population

Of the over 750 participants enrolled in the baseline Smart Work & Life study, 662 participants were included in the final study population (Supplementary Figure 1). Of these study participants, 477 (72%) were female, 36.1% were aged between 35 and 45, 49.2% had a Normal/Underweight body mass index (BMI < 25) and 60.6% were recruited from Leicester. The majority (57.6%) of participants recruited from Leicester were monitored during summer, whereas the majority from Salford (69.7%) were in autumn. On average, participants provided 7.7 days of valid activPAL accelerometry data, with approximately 8.8 hours a day spent sedentary, broken into on average 6.2 prolonged bouts lasting at least 30 minutes, making up 73.0% of the detected sedentary time (Table 1).

**Table 1:** Characteristics of the study population.

| Characteristic | Overall | Leicester | Salford |
| --- | --- | --- | --- |
| **n (%)** | 662 | 401 (60.6) | 261 (39.4) |
| **Age group, n (%)** |  |  |  |
| Under 35 | 122 (18.4) | 84 (20.9) | 38 (14.6) |
| 35 to 45 | 170 (25.7) | 95 (23.7) | 75 (28.7) |
| 45 to 55 | 239 (36.1) | 129 (32.2) | 110 (42.1) |
| 55 and over | 131 (19.8) | 93 (23.2) | 38 (14.6) |
| **Sex, n (%)** |  |  |  |
| Female | 477 (72.1) | 281 (70.1) | 196 (75.1) |
| Male | 185 (27.9) | 120 (29.9) | 65 (24.9) |
| **BMI category, n (%)** |  |  |  |
| Normal/Underweight ($<25 \text{ kgm}^{-2}$) | 326 (49.2) | 196 (48.9) | 130 (49.8) |
| Overweight ($25 - 30 \text{ kgm}^{-2}$) | 195 (29.5) | 117 (29.2) | 78 (29.9) |
| Obese ($\geq 30 \text{ kgm}^{-2}$) | 141 (21.3) | 88 (21.9) | 53 (20.3) |
| **Season of device wear, n (%)** |  |  |  |
| Spring | 63 (9.5) | 63 (15.7) | 0 (0.0) |
| Summer | 281 (42.4) | 231 (57.6) | 50 (19.2) |
| Autumn | 183 (27.6) | 1 (0.2) | 182 (69.7) |
| Winter | 135 (20.4) | 106 (26.4) | 29 (11.1) |
| **Reference measure (activPAL), mean (SD)** |  |  |  |
| Daily sedentary time [hours/day] | 8.8 (2.0) | 9.0 (1.9) | 8.5 (2.1) |
| Daily prolonged sedentary bouts count [number/day] | 6.2 (1.2) | 6.2 (1.2) | 6.1 (1.1) |
| Proportion of sedentary time in prolonged bouts [%] | 73.0 (8.8) | 73.3 (8.8) | 72.5 (8.8) |
| Valid monitoring duration [days] | 7.7 (1.4) | 7.9 (1.2) | 7.6 (1.6) |

BMI - body mass index,
SD – standard deviation, n – population count.
Prolonged sedentary bouts – periods of detected sedentary time lasting $\geq 30$ minutes, allowing brief non-sedentary interruptions ( $\leq 90$ s), with $\geq 80\%$ of the period classified as sedentary.

### Epoch-Level Agreement

The wrist-worn accelerometer models were compared in classification outputs on a 30-second epoch level, to the reference thigh data. Table 2 presents the averages for per-participant performance for the six models evaluated in this analysis. Across three of the four evaluation metrics, OxWearables’ ActiNet model had the highest performance, with an accuracy of 0.87 ± 0.05, binary F1 score of 0.85 ± 0.06, and Cohen’s kappa of 0.73 ± 0.09. However, GGIR default had the highest per-participant sensitivity with 0.91 ± 0.05. The Actimetric Trost extended model has the worst average performance across most evaluation metrics (accuracy 0.73, F1 0.74, Cohen’s kappa 0.48). Per-participant performance distributions (Supplementary Figure S2) supported this ranking: ActiNet achieved the highest epoch-level performance across accuracy, F1, and Cohen’s kappa, followed by the OxWearables Accelerometer model and the three GGIR-based models, which performed comparably to one another. Actimetric Trost was the lowest-performing model overall.

**Table 2:** Epoch-level sedentary classification performance of six wrist-worn accelerometer models relative to a thigh-worn algorithm as reference.

| Model | Accuracy | F1 | Cohen's Kappa | Sensitivity | Specificity |
| --- | --- | --- | --- | --- | --- |
| OxWearables - Accelerometer | 0.843 ± 0.048 | 0.823 ± 0.057 | 0.680 ± 0.095 | 0.874 ± 0.061 | 0.819 ± 0.071 |
| OxWearables - ActiNet | **0.869 ± 0.047** | **0.850 ± 0.055** | **0.733 ± 0.094** | 0.885 ± 0.057 | **0.857 ± 0.065** |
| Actimetric - Trost | 0.733 ± 0.060 | 0.735 ± 0.065 | 0.479 ± 0.107 | 0.882 ± 0.056 | 0.622 ± 0.092 |
| GGIR - Bakrania ENMO (32.6mg) | 0.799 ± 0.050 | 0.785 ± 0.061 | 0.597 ± 0.099 | 0.877 ± 0.062 | 0.741 ± 0.075 |
| GGIR - default ENMO (40mg) | 0.795 ± 0.051 | 0.788 ± 0.059 | 0.594 ± 0.098 | **0.911 ± 0.052** | 0.709 ± 0.077 |
| GGIR - Fraysse ENMOabs (62.5mg) | 0.797 ± 0.050 | 0.788 ± 0.059 | 0.597 ± 0.098 | 0.896 ± 0.055 | 0.724 ± 0.077 |

Performance scores are reported as mean ± standard deviation across all participants in the study. Highest scoring model for respective performance metric is indicated in bold.

### Participant-Level Agreement

We compared the agreement in extracted digital measures (features) from each classification model. Equivalence bounds (±10%) for the accuracy of each algorithm relative to the thigh worn reference are displayed in Figure 2(a)-(c), with exact values reported in Supplementary tables 2(a)-(c). All six wrist-based algorithms overestimated the mean daily sedentary time relative to the thigh-worn reference of 8.8 hours/day. Only the OxWearables ActiNet model fell within the 10% equivalency region for this measure, with mean (90% confidence interval (CI)) of 0.75 (0.68 - 0.82) hours. ActiNet also demonstrated the smallest error (RMSE and MAE) and highest correlation with the reference data.

**Figure 1.**
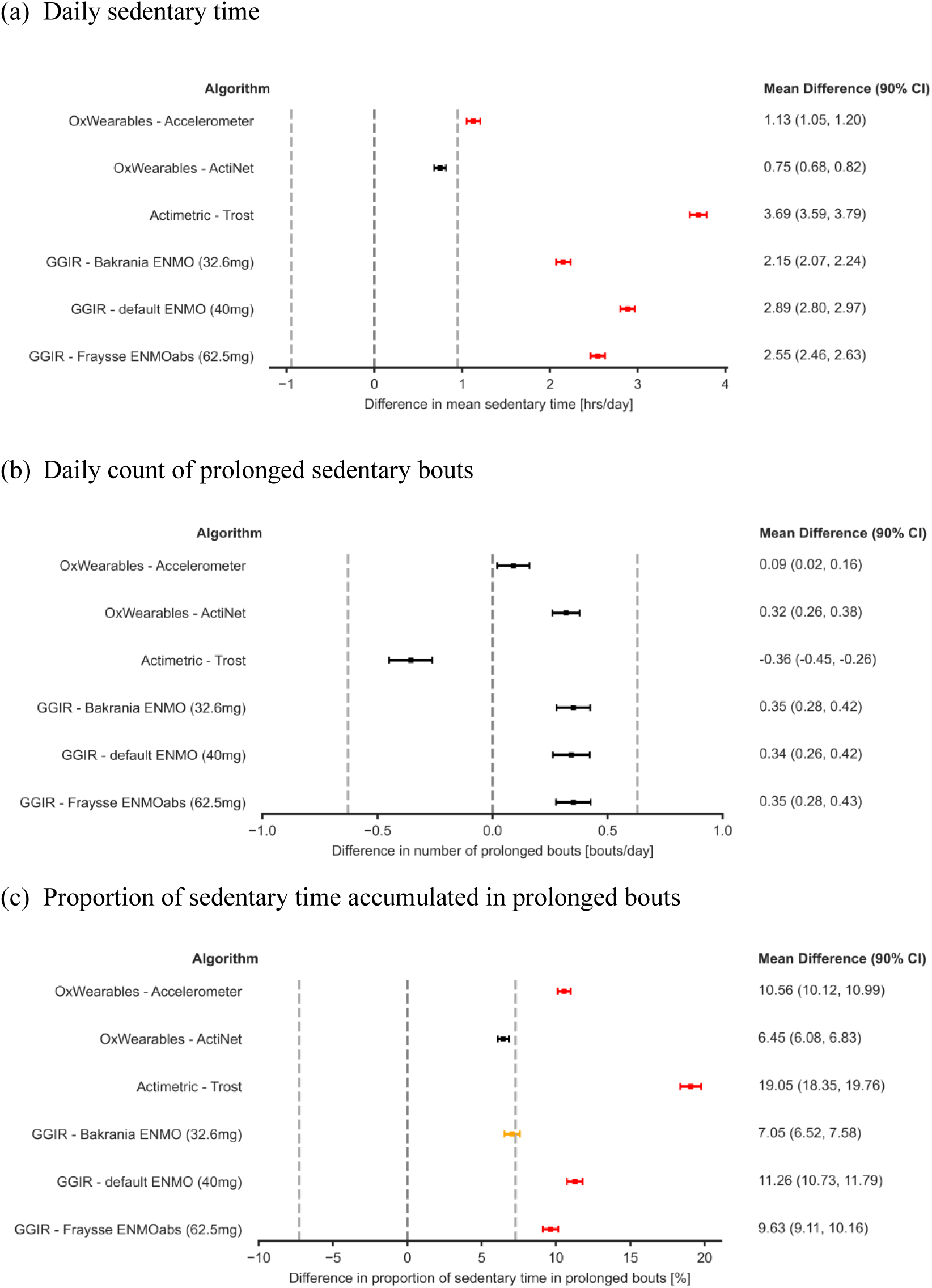
**(a)-(c):** Equivalency of wrist-derived sedentary metrics across six wrist-worn accelerometer models relative to a thigh-worn reference (PALbatch CREA algorithm). Wrist models were compared to the thigh monitor PALbatch CREA algorithm reference. Values within ±10% of the mean reference value, indicated with the light grey dashed line are considered equivalent. Black - equivalent, Yellow - borderline equivalent, Red - not equivalent.

**Figure 2:**
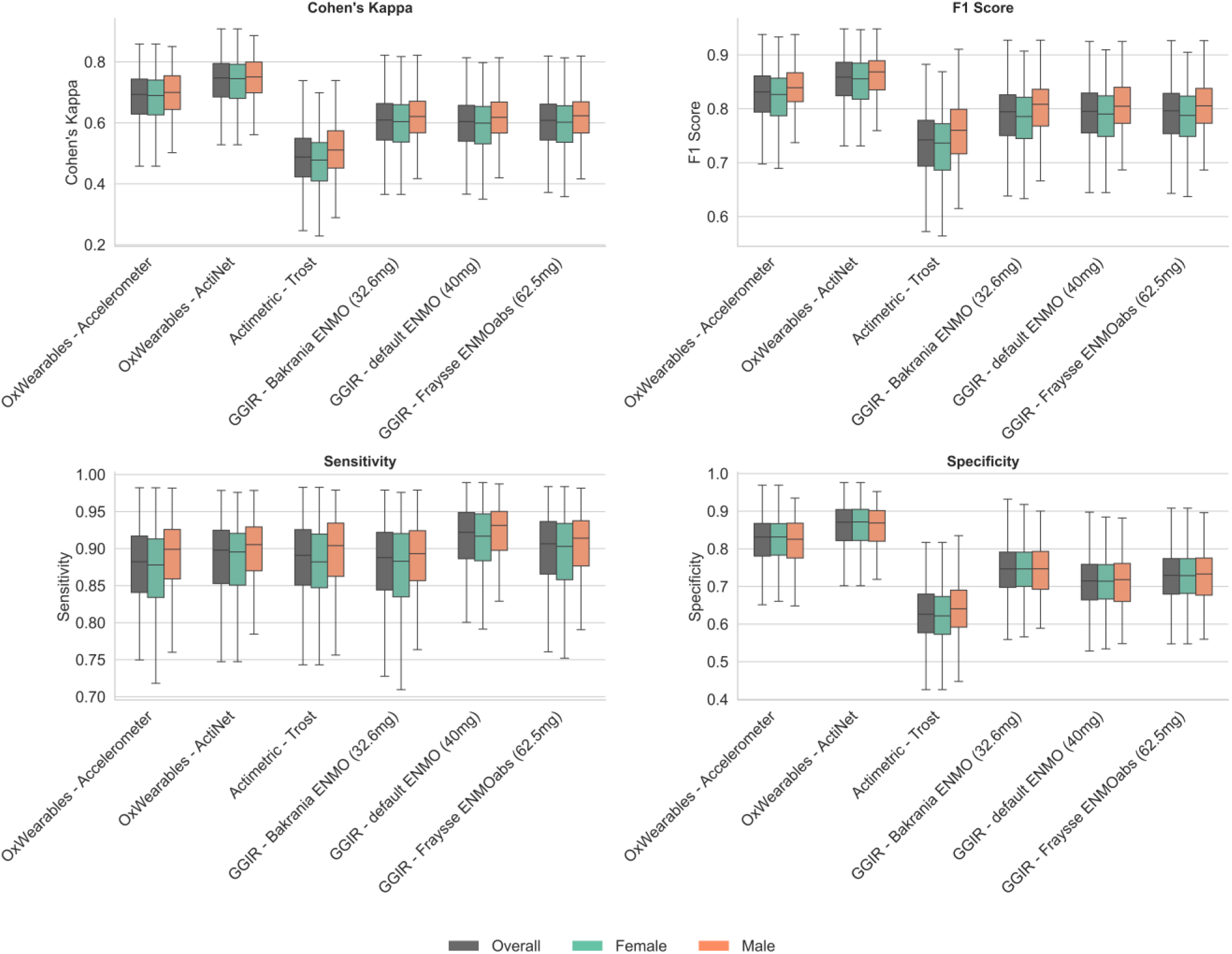
Distribution of epoch-level sedentary classification performance across six wrist-worn accelerometers relative to a thigh worn reference, stratified by sex. Wrist worn models compared to thigh PALbatch CREA algorithm reference.

**Figure 3(a)–(c):**
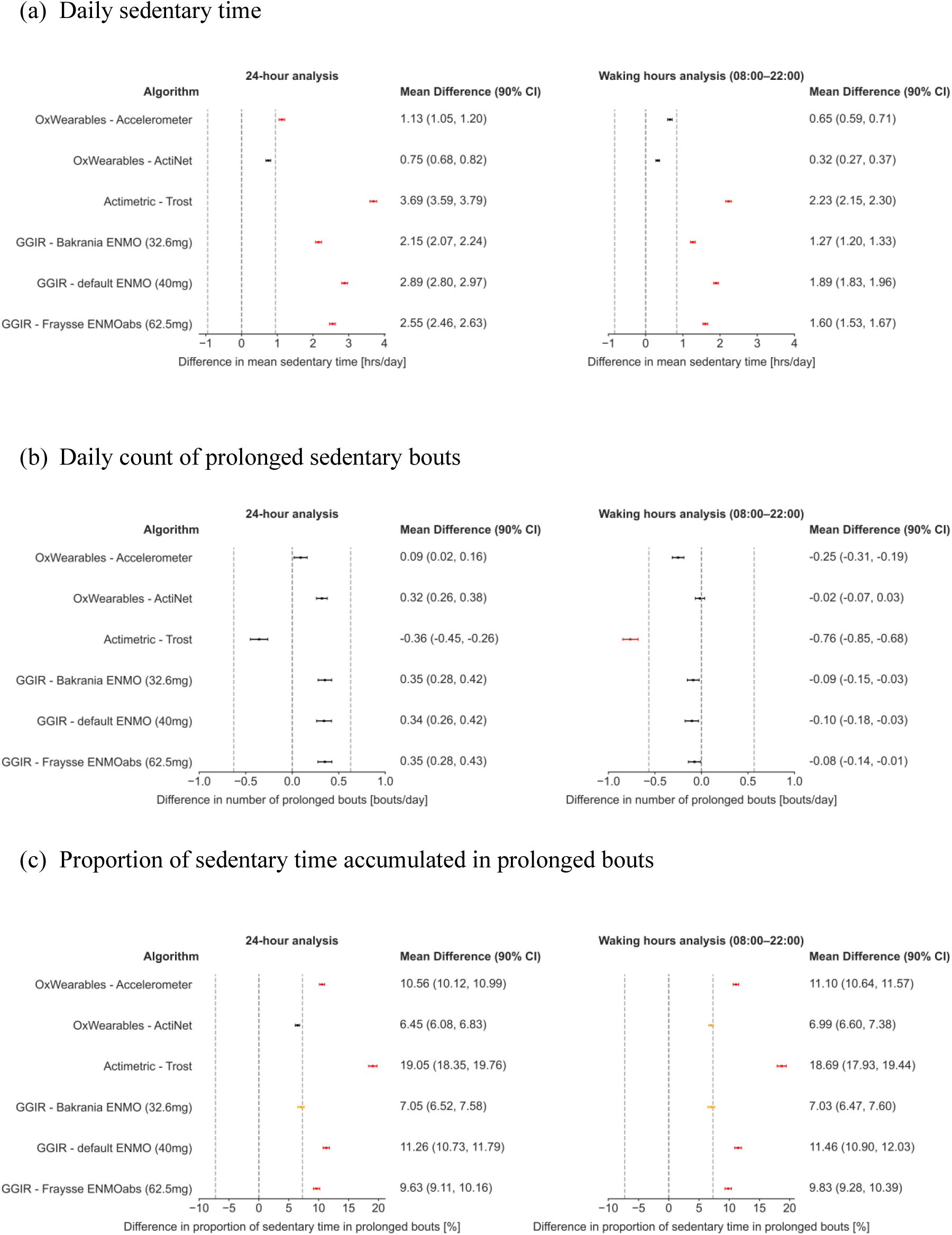
Equivalency of wrist-derived sedentary metrics across six wrist-worn accelerometer models relative to a thigh-worn reference (PALbatch CREA algorithm), restricted to waking hours (08:00–22:00). Wrist models were compared to the thigh monitor PALbatch CREA algorithm reference. Values within ±10% of the mean reference value, indicated with the light grey dashed line are considered equivalent. Black - equivalent, Yellow - borderline equivalent, Red - not equivalent.

For mean daily number of prolonged sedentary bouts, all six algorithms produced estimates within the equivalency region. Although the Accelerometer model produced mean estimates closest to the reference values on average, ActiNet achieved the lowest errors and highest correlation.

For the proportion of sedentary time spent in prolonged bouts, only ActiNet produced mean estimates and 90% CI that fell entirely within the equivalency bounds. As detailed in Supplementary Tables 2(a)-(c), ActiNet demonstrated strong agreement across all three sedentary features, with the lowest RMSE and MAE and the highest Pearson correlation coefficient.

### Subgroup Analysis

To assess the robustness of these findings, we examined model performance across five subgroup categories: age group, sex, BMI group, recruitment centre, and season of wear. Model rankings were broadly preserved between sexes, with ActiNet consistently achieving the highest performance (Figure 2). The same pattern was observed across age, BMI, recruitment centre, and season of wear (Supplementary Tables 3-7; Supplementary Figures 5-8). Performance differences between models therefore, appear robust to variation in participant characteristics and data collection conditions.

### Sensitivity Analysis

We conducted sensitivity analyses restricting the analysis window to waking hours (8:00-22:00) to assess whether model performance and feature agreement were influenced by overnight data.

Restricting to waking hours produced modest changes in epoch-level performance. For the best performing model (OxWearables ActiNet), the per-participant F1 score improved slightly to 0.88 ± 0.05, (up from 0.85 ± 0.06 in the main analysis). The Cohen’s kappa score, however, decreased to 0.68 ± 0.12 in the sensitivity analysis (from 0.73 ± 0.09. (Supplementary Table 8 and Supplementary Figure 8).

The waking hours restriction also influenced agreement in extracted digital measures, (Figure 4(a)-(c)). For the mean daily sedentary time, both OxWearables – Accelerometer and ActiNet models fell within the equivalency bounds in the sensitivity analysis, compared to ActiNet alone in the main analysis. All models trended closer to the reference value with the removal of nighttime windows, due to the removal of wrist models’ false labelling of sleep behaviour as sedentary. The sensitivity analysis also had lower RMSE and MAE errors and higher correlation to the reference, as compared to the main analysis.

For the average daily number of prolonged sedentary bouts, only the Actimetric Trost model was found to be not equivalent. For the ActiNet model, the removal of night time windows produced a near perfect agreement in the daily average number of prolonged sedentary bouts, with a mean (90% CI) difference of −0.02 (−0.07, 0.03).

The proportion of sedentary time in prolonged bouts, however, was more poorly estimated after the removal of nighttime windows. Only the ActiNet and GGIR Bakrania models were found to be borderline equivalent, with respective mean values of 6.99 (6.60, 7.38) and 7.03 (6.47, 7.60), within the equivalence band, yet upper limits (90% CI) greater than 10% of the mean reference value (7.30).

## Discussion

This study is the largest single evaluation of wrist-worn accelerometers for measuring sedentary behaviour. To our knowledge it is also the first to assess equivalency in accelerometer-derived sedentary metrics beyond total daily sedentary time. We compared six models against a thigh-worn reference in 662 working-age adults monitored under free-living conditions for a mean of 7.7 days. Performance varied substantially across models. ActiNet, a self-supervised deep learning model, consistently outperformed the others, achieving the highest epoch-level classification accuracy and the strongest agreement across all derived sedentary metrics. The remaining models showed weaker and more inconsistent performance, with most failing to demonstrate equivalency in estimated daily sedentary time. The number of prolonged sedentary bouts was estimated with reasonable accuracy across wrist-based models, whereas the proportion of sedentary time accumulated in prolonged bouts was less reliably captured.

ActiNet’s strong performance was robust across key demographic subgroups and when analyses were restricted to daytime hours (08:00–22:00), suggesting it may offer a viable alternative to thigh-worn devices for sedentary behaviour assessment in similar populations. The remaining five models performed comparatively worse across both classification and metric agreement, in contrast to prior work that has typically reported high agreement and strong correlations^26,50^. For example, Matthews et al. (2025) ^26^, reported good agreement and low root mean squared errors for daily sedentary time using five wrist-based models against a thigh monitor during daytime hours in 71 US-based adults aged 30 to 65. In the present study, four of these five models did not demonstrate equivalency in estimated daytime sedentary time, with only the Ox Wearables Accelerometer model achieving equivalency. This discrepancy may partly reflect the substantially larger sample in the present study (n = 662 vs. n = 71), which likely provides a more robust characterisation of model performance under free-living conditions.

Models employing more sophisticated learning frameworks, larger and more representative training datasets, and temporal smoothing of predictions consistently demonstrated stronger performance. Within the traditional ML models, the contrast between the Accelerometer and Trost models may be informative to future directions in this space. The Trost model employs a random forest classifier with 100 trees, trained on just 16 Australian participants, whereas the Accelerometer model uses a random forest with 200 trees, trained on 151 UK participants^34,38^. In addition, the Accelerometer model incorporates hidden Markov model smoothing and post hoc sleep correction, enabling it to account for temporal dependencies in activity patterns. The absence of such smoothing in the Trost model likely contributed to misclassification of brief activity interruptions as sedentary time, resulting in systematic overestimation of sedentary behaviour and explaining its underperformance relative to earlier studies^26,38,40^. Consistent with previous studies we found significance performance limitations across a range of cut point thresholds, highlighting ongoing challenges in selecting appropriate parameter values^42,50,51^. This sensitivity to threshold choice, and the limited adaptability of cut point-based approaches across populations and contexts, represents a fundamental methodological limitation. Even when implemented within widely used processing frameworks such as GGIR, careful consideration of cut point selection remains essential for valid sedentary behaviour estimation.

Our findings shed a first light on the reliability of more complex bout-level metrics. While daily prolonged sedentary bout number could be estimated with reasonable accuracy across wrist-based models, the proportion of sedentary time accumulated in prolonged bouts was less reliably captured. This suggests that features reflecting sedentary accumulation patterns are more sensitive to model limitations than simpler count-based metrics, and points to an important area for future model development as the field moves towards more nuanced characterisation of sedentary behaviour.

Our study has numerous strengths. These include a relatively large sample of 662 participants, with an average monitoring duration exceeding one week, providing a comprehensive dataset for the external validation of six sedentary behaviour classification models. The study population was well characterised, with detailed demographic information enabling exploratory subgroup analyses by age, sex, BMI, and other relevant factors. Data were collected under free-living conditions, enhancing the validity of the findings and ensuring that results reflect typical real-world patterns of behaviour. A further strength is the comprehensive evaluation framework, with models assessed at both the epoch and participant levels, incorporating multiple sedentary-related metrics. This allows for a more complete comparison of model performance, particularly for applications where derived features are of primary interest. Sensitivity analyses examining both 24-hour and daytime (08:00-22:00) periods further strengthen the findings by demonstrating robustness independent of sleep detection methods. Finally, the use of open-source code and models, on the SMART Work & Life dataset that is available upon request, supports transparency and reproducibility of these analyses.

There are also limitations to consider. First, the study population comprised working-age office-based adults recruited from England. As such, participants may not exhibit the full range of behaviours representative of the wider UK or global populations, potentially limiting generalisability. Although thigh-worn accelerometers are widely accepted as a reference standard, they may have limited ability to distinguish between primary (sleep-related) and secondary (waking) lying postures. Additionally, a key challenge in accelerometer-based studies is distinguishing sedentary behaviour from device non-wear. In this study, wrist non-wear was difficult to identify when a participant continued to wear the thigh monitor. This means some non-wear time was likely misclassified as sedentary behaviour, potentially inflating wrist-based estimates.. To enable consistent comparison, outputs were standardised to the lowest common frequency of window labels every 30 seconds. This approach limited the ability to evaluate epoch-level performance at finer temporal resolutions. Additionally, the identification of prolonged sedentary bouts allowed for interruptions of up to 90 seconds, reflecting three consecutive, non-sedentary, 30-second windows. This resulted in a coarser characterisation of prolonged sedentary bouts, overlooking shorter, more transient interruptions in sedentary behaviour.

In conclusion, this study shows that wrist-worn accelerometers can provide accurate measurements of sedentary behaviour in free-living settings. Substantial performance variability across the evaluated models underscores that model selection is a critical determinant of measurement accuracy in wrist-worn accelerometery. While some models produced reasonable estimates of total sedentary time, fewer achieved equivalency, and performance was generally weaker for more complex measurements such as the proportion of sedentary time accumulated in prolonged bouts. This work provides confidence in future epidemiological research to examine sedentary behaviour patterns from wrist-worn accelerometers and their associations with health outcomes.

## Declarations

### Funding

The SMART Work & Life study was funded by the National Institute for Health and Care Research (NIHR) public health research programme [PR-R5-0213-25004].

### Data Availability

This study uses the SMART Work & Life dataset, which is available from the corresponding author, BM, upon reasonable request.

### Code Availability

The code for this study is available on Github and can be accessed via this link: https://github.com/OxWearables/sedentary-detection.

## Notes

### Competing Interest Statement

AD has released the OxWearables - Accelerometer package under an academic use licence that has resulted in commercial entities paying license fees for their use.

### Author Declarations

The SMART Work & Life study received ethical approval from the University of Leicester's Sub-Committee for Medicine and Biological Sciences (Ref: 14372) and the University of Salford's Research Enterprise and Engagement ethical approval panel (Ref: HSR1718-039) prior to the commencement of the study. All participants provided written informed consent before taking part.

