## Supplementary Materials for "Evaluating Open-Source Wrist-Worn Accelerometer Models for Sedentary Time Detection Against Thigh-Worn Accelerometer Data"

### Study Population

Supplementary Figure 1: Participant exclusion flow diagram.

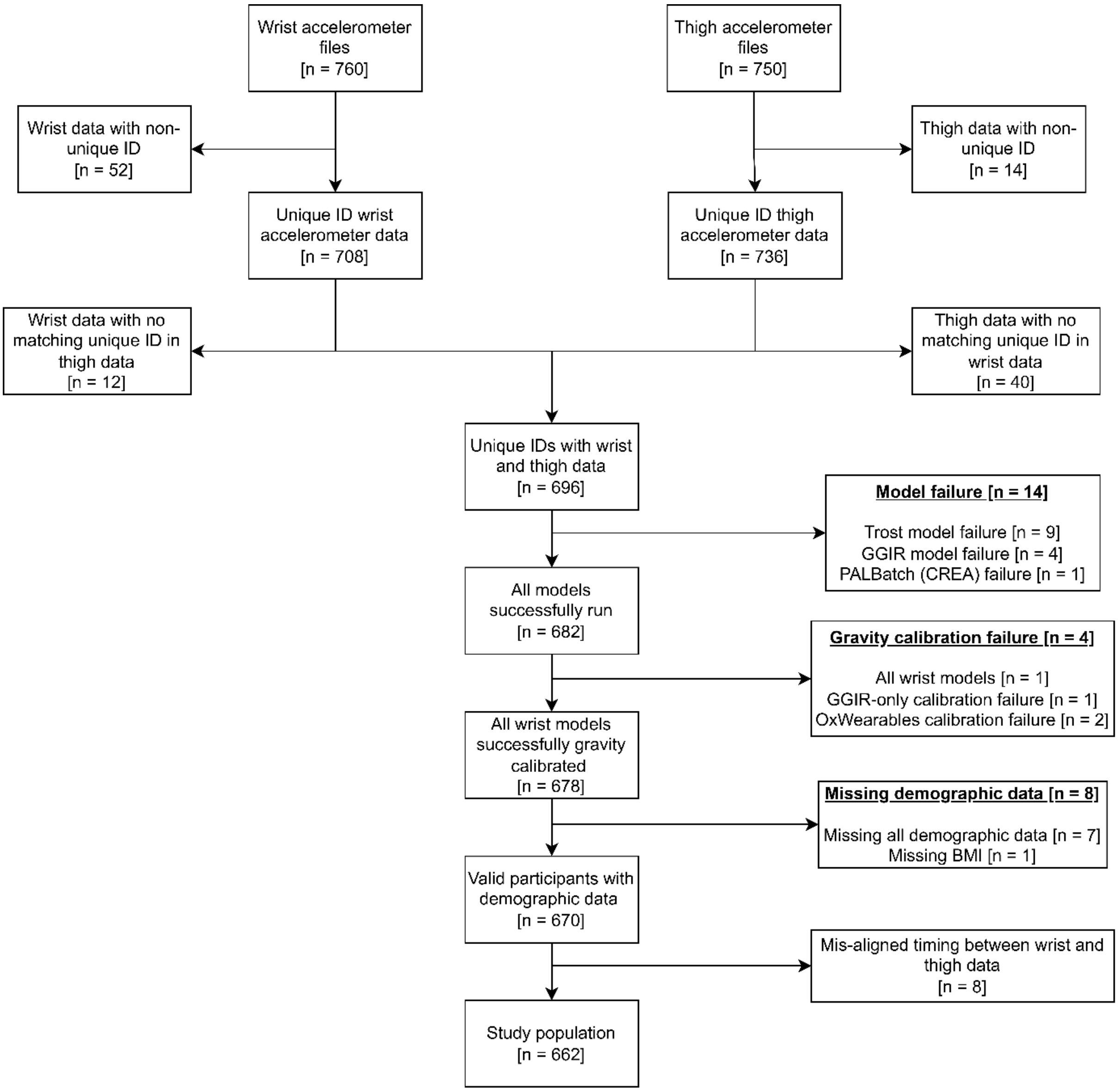

Supplementary Figure 2: Distribution of epoch-level sedentary classification performance across six wrist-worn accelerometer models, relative to a thigh-worn algorithm as reference.

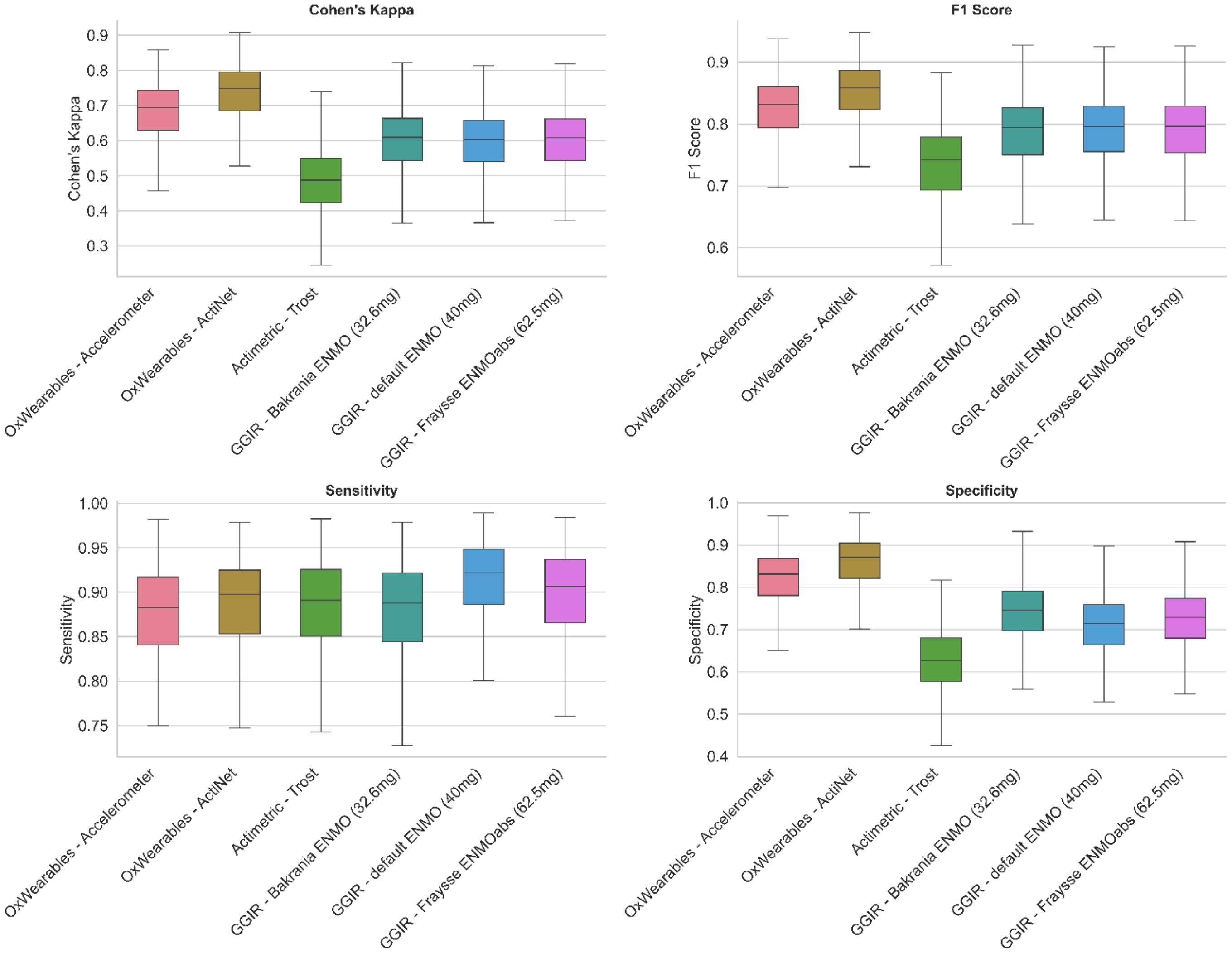

Supplementary Figure 3: Representative sample plot from a single participant and day.

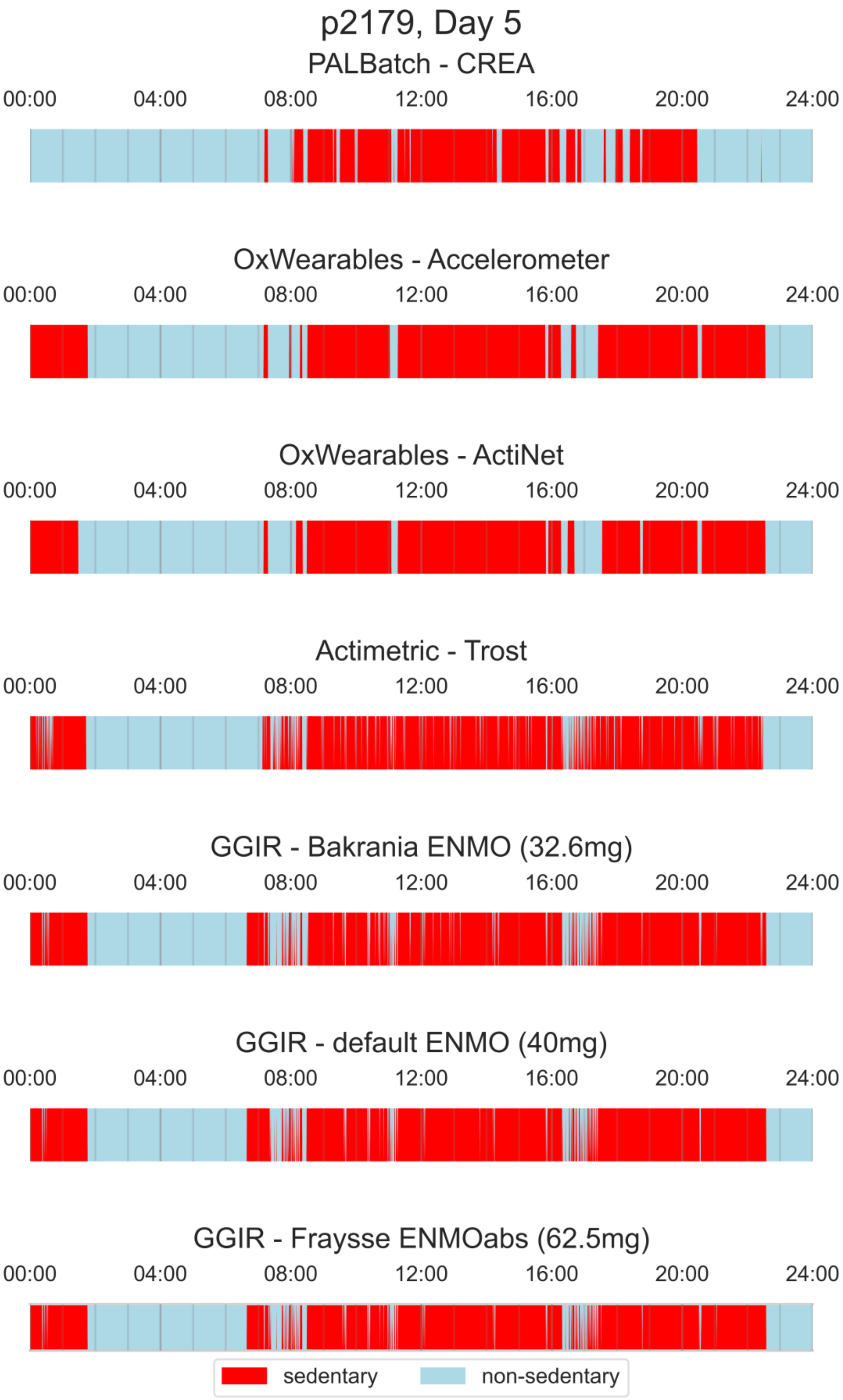

### Epoch-level Agreement

**Supplementary Figure 4:** Confusion matrices for epoch-level sedentary classification across six wrist-worn accelerometer models relative to a thigh-worn reference, based on 30-second windows.

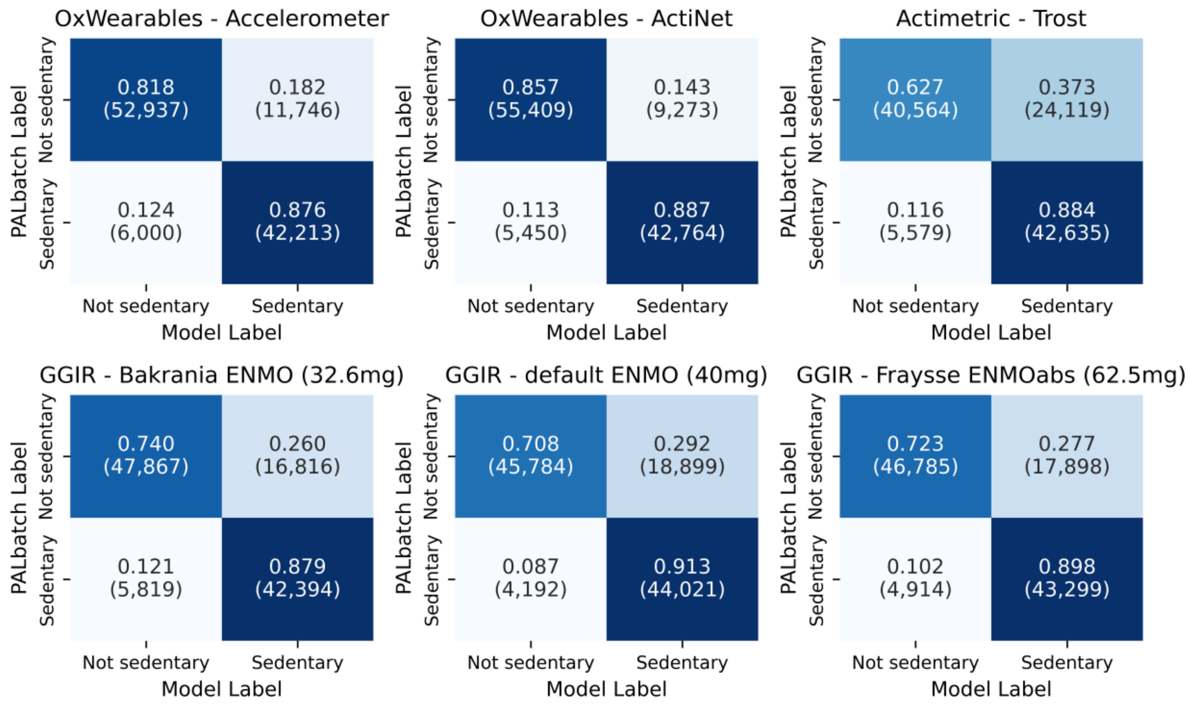

Confusion matrices are aggregated across all valid participant windows, with the number of hours for each cell in parentheses.

**Supplementary Table 1(a)–(f):** Cross-tabulations of the most frequent 30-second epoch label across six wrist-worn accelerometer models relative to a thigh-worn reference (PALbatch CREA algorithm).

#### OxWearables - Accelerometer

| palbatch | sleep | sedentary | light | moderate-vigorous |
|---|---|---|---|---|
| **sleep** | 0.88<br>(2,021,438) | 0.11<br>(263,622) | 0.00<br>(10,990) | 0.00<br>(227) |
| **lying** | 0.25<br>(31,017) | 0.73<br>(90,020) | 0.02<br>(2,357) | 0.00<br>(112) |
| **sedentary** | 0.02<br>(49,397) | 0.91<br>(2,247,262) | 0.07<br>(170,823) | 0.00<br>(6,215) |
| **vehicle** | 0.00<br>(14) | 0.66<br>(195,586) | 0.33<br>(97,356) | 0.01<br>(2,799) |
| **upright** | 0.00<br>(5,575) | 0.31<br>(429,308) | 0.65<br>(895,178) | 0.03<br>(46,446) |
| **walking** | 0.00<br>(68) | 0.05<br>(10,260) | 0.32<br>(64,256) | 0.62<br>(123,142) |
| **bicycling** | 0.00<br>(1) | 0.15<br>(1,509) | 0.12<br>(1,236) | 0.73<br>(7,567) |

#### OxWearables - ActiNet

| palbatch | sleep | sedentary | light | moderate-vigorous |
|---|---|---|---|---|
| **sleep** | 0.88<br>(2,029,901) | 0.11<br>(259,144) | 0.00<br>(6,924) | 0.00<br>(309) |
| **lying** | 0.25<br>(30,354) | 0.74<br>(90,984) | 0.02<br>(2,020) | 0.00<br>(148) |
| **sedentary** | 0.02<br>(45,170) | 0.90<br>(2,231,025) | 0.08<br>(189,098) | 0.00<br>(8,404) |
| **vehicle** | 0.00<br>(16) | 0.82<br>(243,863) | 0.16<br>(46,958) | 0.02<br>(4,917) |
| **upright** | 0.00<br>(5,932) | 0.21<br>(289,704) | 0.73<br>(1,009,454) | 0.05<br>(71,417) |
| **walking** | 0.00<br>(36) | 0.04<br>(7,132) | 0.28<br>(54,444) | 0.69<br>(136,114) |
| **bicycling** | 0.00<br>(0) | 0.04<br>(368) | 0.09<br>(946) | 0.87<br>(8,998) |

#### Actimetric - Trost

| palbatch | nighttime.sleep | nighttime.awake | sedentary | stationary | walk | run |
|---|---|---|---|---|---|---|
| **sleep** | 0.72<br>(1,641,851) | 0.09<br>(202,544) | 0.19<br>(443,898) | 0.00<br>(4,705) | 0.00<br>(3,128) | 0.00<br>(152) |
| **lying** | 0.05<br>(6,086) | 0.01<br>(1,198) | 0.92<br>(113,172) | 0.01<br>(1,542) | 0.01<br>(1,460) | 0.00<br>(48) |
| **sedentary** | 0.01<br>(23,595) | 0.00<br>(8,487) | 0.93<br>(2,299,010) | 0.03<br>(67,474) | 0.03<br>(73,425) | 0.00<br>(1,706) |
| **vehicle** | 0.00<br>(0) | 0.00<br>(39) | 0.62<br>(182,802) | 0.36<br>(105,135) | 0.03<br>(7,428) | 0.00<br>(350) |
| **upright** | 0.00<br>(3,080) | 0.00<br>(6,192) | 0.73<br>(1,010,645) | 0.12<br>(171,412) | 0.13<br>(175,815) | 0.01<br>(9,362) |
| **walking** | 0.00<br>(10) | 0.00<br>(150) | 0.39<br>(77,494) | 0.06<br>(11,312) | 0.50<br>(97,972) | 0.05<br>(10,787) |
| **bicycling** | 0.00<br>(0) | 0.00<br>(4) | 0.20<br>(2,046) | 0.40<br>(4,163) | 0.26<br>(2,730) | 0.13<br>(1,370) |

#### GGIR - Bakrania ENMO (32.6mg)

| palbatch | sleep | inactive | light | moderate-vigorous |
|---|---|---|---|---|
| **sleep** | 0.84<br>(1,937,151) | 0.15<br>(352,620) | 0.00<br>(4,980) | 0.00<br>(1,526) |
| **lying** | 0.09<br>(10,776) | 0.89<br>(110,324) | 0.02<br>(1,896) | 0.00<br>(511) |
| **sedentary** | 0.01<br>(32,596) | 0.92<br>(2,272,895) | 0.06<br>(138,809) | 0.01<br>(29,396) |
| **vehicle** | 0.00<br>(597) | 0.57<br>(169,318) | 0.39<br>(114,166) | 0.04<br>(11,674) |
| **upright** | 0.01<br>(12,424) | 0.48<br>(660,956) | 0.38<br>(524,832) | 0.13<br>(178,296) |
| **walking** | 0.00<br>(509) | 0.07<br>(13,366) | 0.21<br>(41,222) | 0.72<br>(142,630) |
| **bicycling** | 0.00<br>(28) | 0.17<br>(1,770) | 0.21<br>(2,201) | 0.61<br>(6,314) |

#### GGIR - default ENMO (40mg)

| palbatch | sleep | inactive | light | moderate-vigorous |
|---|---|---|---|---|
| **sleep** | 0.84<br>(1,937,124) | 0.15<br>(354,750) | 0.00<br>(2,958) | 0.00<br>(1,446) |
| **lying** | 0.09<br>(10,775) | 0.90<br>(111,186) | 0.01<br>(1,064) | 0.00<br>(482) |
| **sedentary** | 0.01<br>(32,594) | 0.94<br>(2,327,004) | 0.03<br>(84,738) | 0.01<br>(29,360) |
| **vehicle** | 0.00<br>(596) | 0.72<br>(212,559) | 0.24<br>(70,046) | 0.04<br>(12,554) |
| **upright** | 0.01<br>(12,423) | 0.57<br>(784,156) | 0.29<br>(396,012) | 0.13<br>(183,916) |
| **walking** | 0.00<br>(509) | 0.09<br>(17,672) | 0.18<br>(36,434) | 0.72<br>(143,112) |
| **bicycling** | 0.00<br>(28) | 0.20<br>(2,034) | 0.19<br>(1,908) | 0.62<br>(6,343) |

#### GGIR - Fraysse ENMOabs (62.5mg)

| palbatch | sleep | inactive | light | moderate-vigorous |
|---|---|---|---|---|
| **sleep** | 0.84<br>(1,937,132) | 0.15<br>(354,003) | 0.00<br>(1,134) | 0.00<br>(4,008) |
| **lying** | 0.09<br>(10,775) | 0.90<br>(110,887) | 0.00<br>(492) | 0.01<br>(1,352) |
| **sedentary** | 0.01<br>(32,594) | 0.93<br>(2,311,599) | 0.02<br>(37,306) | 0.04<br>(92,197) |
| **vehicle** | 0.00<br>(596) | 0.66<br>(196,186) | 0.18<br>(53,618) | 0.15<br>(45,355) |
| **upright** | 0.01<br>(12,422) | 0.54<br>(743,462) | 0.09<br>(127,982) | 0.36<br>(492,641) |
| **walking** | 0.00<br>(509) | 0.07<br>(14,490) | 0.03<br>(5,697) | 0.90<br>(177,030) |
| **bicycling** | 0.00<br>(28) | 0.18<br>(1,888) | 0.05<br>(474) | 0.77<br>(7,924) |

| | Mean (SD) | RMSE | MAE | Pearson's r |
|---|---|---|---|---|
| **PALBatch - CREA [ref]** | 9.490 (1.533) | - | - | - |

| | | | | |
|---|---|---|---|---|
| **OxWearables - Accelerometer** | 10.618 (1.733) | 1.656 | 1.295 | 0.730 |
| **OxWearables - ActiNet** | 10.236 (1.683) | 1.298 | 0.967 | 0.785 |
| **Actimetric - Trost** | 13.186 (1.588) | 3.983 | 3.701 | 0.546 |
| **GGIR - Bakrania ENMO (32.6mg)** | 11.645 (1.722) | 2.513 | 2.201 | 0.689 |
| **GGIR - default ENMO (40mg)** | 12.378 (1.710) | 3.165 | 2.899 | 0.686 |
| **GGIR - Fraysse ENMOabs (62.5mg)** | 12.039 (1.713) | 2.858 | 2.568 | 0.687 |

Prolonged sedentary bouts are defined as detected periods of sedentary behaviour, lasting at least 30 minutes, with no gap in sedentary behaviour for over 90 seconds, and minimum proportion of detected sedentary behaviour of 80%.
RMSE – root mean squared error, MAE – mean absolute error, Pearson's r – Pearson's correlation coefficient, SD – standard deviation.

(a) Average daily number of prolonged sedentary bouts (count/day)

| | Mean (SD) | RMSE | MAE | Pearson's r |
|---|---|---|---|---|
| **PALBatch - CREA [ref]** | 6.29 (1.20) | - | - | - |
| **OxWearables - Accelerometer** | 6.38 (1.03) | 1.11 | 0.84 | 0.518 |
| **OxWearables - ActiNet** | 6.61 (1.14) | 0.98 | 0.72 | 0.690 |
| **Actimetric - Trost** | 5.93 (1.05) | 1.51 | 1.17 | 0.149 |
| **GGIR - Bakrania ENMO (32.6mg)** | 6.65 (1.08) | 1.20 | 0.93 | 0.495 |
| **GGIR - default ENMO (40mg)** | 6.64 (1.08) | 1.30 | 1.01 | 0.396 |
| **GGIR - Fraysse ENMOabs (62.5mg)** | 6.65 (1.04) | 1.23 | 0.97 | 0.450 |

Prolonged sedentary bouts are defined as detected periods of sedentary behaviour, lasting at least 30 minutes, with no gap in sedentary behaviour for over 90 seconds, and minimum proportion of detected sedentary behaviour of 80%.
RMSE – root mean squared error, MAE – mean absolute error, Pearson's r – Pearson's correlation coefficient, SD – standard deviation.

(b) Proportion of sedentary time in prolonged sedentary bouts (%)

| | Mean (SD) | RMSE | MAE | Pearson's r |
|---|---|---|---|---|
| **PALBatch - CREA [ref]** | 72.7 (9.0) | - | - | - |
| **OxWearables - Accelerometer** | 83.3 (6.0) | 12.6 | 10.7 | 0.647 |
| **OxWearables - ActiNet** | 79.2 (7.0) | 8.7 | 6.9 | 0.759 |
| **Actimetric - Trost** | 91.8 (9.1) | 22.1 | 19.5 | 0.263 |
| **GGIR - Bakrania ENMO (32.6mg)** | 79.8 (8.4) | 10.9 | 8.5 | 0.554 |
| **GGIR - default ENMO (40mg)** | 84.0 (7.7) | 14.0 | 11.6 | 0.519 |
| **GGIR - Fraysse ENMOabs (62.5mg)** | 82.4 (7.9) | 12.7 | 10.2 | 0.540 |

Prolonged sedentary bouts are defined as detected periods of sedentary behaviour, lasting at least 30 minutes, with no gap in sedentary behaviour for over 90 seconds, and minimum proportion of detected sedentary behaviour of 80%.
RMSE – root mean squared error, MAE – mean absolute error, Pearson's r – Pearson's correlation coefficient, SD – standard deviation.

### Subgroup Analysis

#### Age group

Supplementary Table 3(a)–(d): Epoch-level sedentary classification performance of six wrist-worn accelerometer models relative to a thigh-worn reference (PALbatch CREA algorithm), stratified by age group.

(a) Performance in participants aged Under 35

| Model | Accuracy | F1 | Cohen's Kappa | Sensitivity | Specificity |
|---|---|---|---|---|---|
| **OxWearables - Accelerometer** | 0.836 ± 0.051 | 0.819 ± 0.056 | 0.666 ± 0.106 | 0.861 ± 0.073 | 0.814 ± 0.074 |
| **OxWearables - ActiNet** | 0.857 ± 0.050 | 0.840 ± 0.056 | 0.708 ± 0.105 | 0.871 ± 0.067 | 0.845 ± 0.067 |
| **Actimetric - Trost** | 0.745 ± 0.062 | 0.746 ± 0.065 | 0.495 ± 0.126 | 0.868 ± 0.066 | 0.647 ± 0.097 |
| **GGIR - Bakrania ENMO (32.6mg)** | 0.798 ± 0.055 | 0.785 ± 0.062 | 0.593 ± 0.115 | 0.861 ± 0.071 | 0.746 ± 0.081 |
| **GGIR - default ENMO (40mg)** | 0.799 ± 0.055 | 0.793 ± 0.059 | 0.598 ± 0.114 | 0.897 ± 0.061 | 0.720 ± 0.084 |
| **GGIR - Fraysse ENMOabs (62.5mg)** | 0.799 ± 0.055 | 0.791 ± 0.060 | 0.597 ± 0.114 | 0.880 ± 0.065 | 0.735 ± 0.083 |

Cells represent the mean ± standard deviation in per-participant performance.

(b) Performance in participants aged 35 to 45

| Model | Accuracy | F1 | Cohen's Kappa | Sensitivity | Specificity |
|---|---|---|---|---|---|
| **OxWearables - Accelerometer** | 0.842 ± 0.042 | 0.818 ± 0.052 | 0.677 ± 0.085 | 0.864 ± 0.066 | 0.825 ± 0.064 |
| **OxWearables - ActiNet** | 0.868 ± 0.040 | 0.845 ± 0.050 | 0.728 ± 0.082 | 0.875 ± 0.064 | 0.862 ± 0.056 |
| **Actimetric - Trost** | 0.730 ± 0.047 | 0.727 ± 0.055 | 0.473 ± 0.086 | 0.870 ± 0.060 | 0.631 ± 0.074 |
| **GGIR - Bakrania ENMO (32.6mg)** | 0.799 ± 0.043 | 0.782 ± 0.053 | 0.597 ± 0.085 | 0.873 ± 0.061 | 0.744 ± 0.067 |
| **GGIR - default ENMO (40mg)** | 0.795 ± 0.044 | 0.785 ± 0.051 | 0.593 ± 0.084 | 0.907 ± 0.051 | 0.713 ± 0.070 |
| **GGIR - Fraysse ENMOabs (62.5mg)** | 0.797 ± 0.044 | 0.784 ± 0.052 | 0.596 ± 0.085 | 0.891 ± 0.055 | 0.729 ± 0.070 |

Cells represent the mean ± standard deviation in per-participant performance.

(c) Performance in participants aged 45 to 55

| Model | Accuracy | F1 | Cohen's Kappa | Sensitivity | Specificity |
|---|---|---|---|---|---|
| **OxWearables - Accelerometer** | 0.844 ± 0.051 | 0.824 ± 0.060 | 0.684 ± 0.096 | 0.878 ± 0.049 | 0.820 ± 0.076 |
| **OxWearables - ActiNet** | 0.873 ± 0.049 | 0.854 ± 0.058 | 0.740 ± 0.096 | 0.893 ± 0.044 | 0.859 ± 0.071 |
| **Actimetric - Trost** | 0.728 ± 0.064 | 0.730 ± 0.068 | 0.472 ± 0.109 | 0.886 ± 0.050 | 0.614 ± 0.097 |
| **GGIR - Bakrania ENMO (32.6mg)** | 0.799 ± 0.051 | 0.783 ± 0.064 | 0.599 ± 0.097 | 0.878 ± 0.060 | 0.743 ± 0.072 |
| **GGIR - default ENMO (40mg)** | 0.795 ± 0.051 | 0.786 ± 0.062 | 0.595 ± 0.095 | 0.913 ± 0.049 | 0.710 ± 0.074 |
| **GGIR - Fraysse ENMOabs (62.5mg)** | 0.798 ± 0.051 | 0.786 ± 0.061 | 0.599 ± 0.095 | 0.899 ± 0.049 | 0.725 ± 0.075 |

Cells represent the mean ± standard deviation in per-participant performance.

(d) Performance in participants aged 55 and over

| Model | Accuracy | F1 | Cohen's Kappa | Sensitivity | Specificity |
|---|---|---|---|---|---|
| **OxWearables - Accelerometer** | 0.846 ± 0.046 | 0.831 ± 0.058 | 0.688 ± 0.094 | 0.889 ± 0.058 | 0.814 ± 0.068 |
| **OxWearables - ActiNet** | 0.876 ± 0.044 | 0.861 ± 0.055 | 0.747 ± 0.089 | 0.898 ± 0.054 | 0.860 ± 0.063 |
| **Actimetric - Trost** | 0.735 ± 0.063 | 0.746 ± 0.070 | 0.484 ± 0.109 | 0.905 ± 0.044 | 0.605 ± 0.094 |
| **GGIR - Bakrania ENMO (32.6mg)** | 0.799 ± 0.053 | 0.792 ± 0.062 | 0.600 ± 0.103 | 0.892 ± 0.055 | 0.729 ± 0.083 |
| **GGIR - default ENMO (40mg)** | 0.792 ± 0.056 | 0.792 ± 0.064 | 0.590 ± 0.106 | 0.923 ± 0.048 | 0.693 ± 0.085 |
| **GGIR - Fraysse ENMOabs (62.5mg)** | 0.795 ± 0.055 | 0.792 ± 0.064 | 0.594 ± 0.104 | 0.908 ± 0.051 | 0.709 ± 0.083 |

Cells represent the mean ± standard deviation in per-participant performance.

**Supplementary Figure 5:** Distribution of epoch-level sedentary classification performance across six wrist-worn accelerometers relative to a thigh worn reference, stratified by age

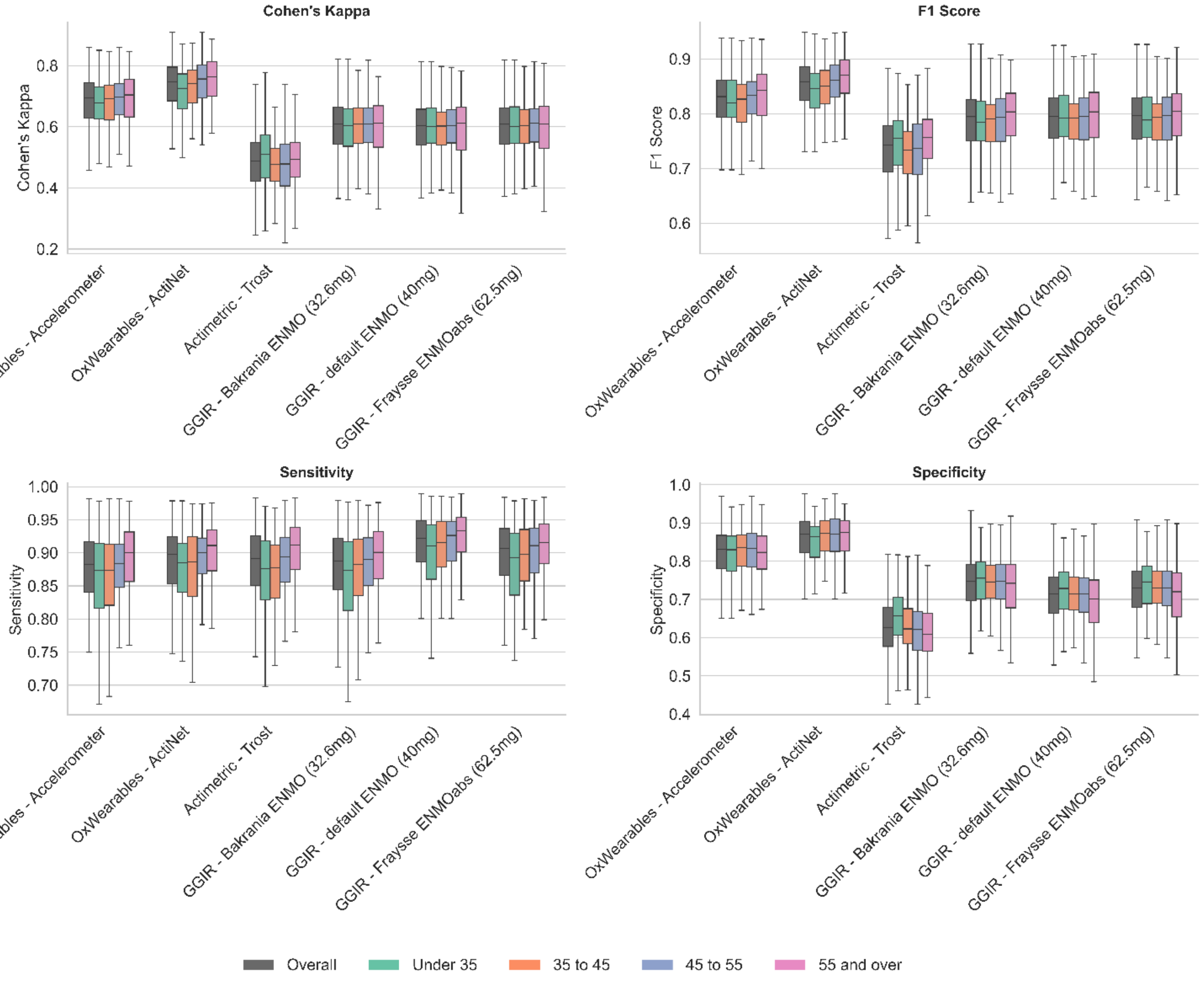

Wrist worn models compared to thigh PALbatch CREA algorithm reference

### Sex

Supplementary Table 4(a)–(b): Epoch-level sedentary classification performance of six wrist-worn accelerometer models relative to a thigh-worn reference (PALbatch CREA algorithm), stratified by sex.

(a) Performance in Female participants

| Model | Accuracy | F1 | Cohen's Kappa | Sensitivity | Specificity |
|---|---|---|---|---|---|
| **OxWearables - Accelerometer** | 0.840 ± 0.050 | 0.818 ± 0.060 | 0.675 ± 0.097 | 0.869 ± 0.061 | 0.820 ± 0.072 |
| **OxWearables - ActiNet** | 0.868 ± 0.048 | 0.846 ± 0.058 | 0.729 ± 0.096 | 0.882 ± 0.057 | 0.857 ± 0.067 |
| **Actimetric - Trost** | 0.726 ± 0.062 | 0.726 ± 0.066 | 0.466 ± 0.108 | 0.878 ± 0.056 | 0.616 ± 0.096 |
| **GGIR - Bakrania ENMO (32.6mg)** | 0.796 ± 0.051 | 0.780 ± 0.063 | 0.592 ± 0.099 | 0.872 ± 0.063 | 0.742 ± 0.074 |
| **GGIR - default ENMO (40mg)** | 0.792 ± 0.052 | 0.783 ± 0.061 | 0.589 ± 0.098 | 0.907 ± 0.053 | 0.709 ± 0.077 |
| **GGIR - Fraysse ENMOabs (62.5mg)** | 0.795 ± 0.051 | 0.782 ± 0.061 | 0.592 ± 0.097 | 0.892 ± 0.055 | 0.7250.077 |

Values represent mean ± standard deviation of per-participant performance.

(b) Performance in Male participants

| Model | Accuracy | F1 | Cohen's Kappa | Sensitivity | Specificity |
|---|---|---|---|---|---|
| **OxWearables - Accelerometer** | 0.848 ± 0.042 | 0.836 ± 0.047 | 0.692 ± 0.088 | 0.885 ± 0.058 | 0.817 ± 0.068 |
| **OxWearables - ActiNet** | 0.874 ± 0.042 | 0.861 ± 0.046 | 0.743 ± 0.087 | 0.893 ± 0.056 | 0.857 ± 0.062 |
| **Actimetric - Trost** | 0.752 ± 0.050 | 0.758 ± 0.056 | 0.511 ± 0.098 | 0.893 ± 0.055 | 0.640 ± 0.080 |
| **GGIR - Bakrania ENMO (32.6mg)** | 0.805 ± 0.048 | 0.799 ± 0.052 | 0.610 ± 0.099 | 0.887 ± 0.058 | 0.740 ± 0.077 |
| **GGIR - default ENMO (40mg)** | 0.802 ± 0.048 | 0.802 ± 0.051 | 0.607 ± 0.098 | 0.919 ± 0.050 | 0.710 ± 0.079 |
| **GGIR - Fraysse ENMOabs (62.5mg)** | 0.804 ± 0.048 | 0.801 ± 0.052 | 0.610 ± 0.099 | 0.904 ± 0.053 | 0.725 ± 0.078 |

Values represent mean ± standard deviation of per-participant performance.

### BMI category

Supplementary Table 5(a)-(c): Epoch-level sedentary classification performance of six wrist-worn accelerometer models relative to a thigh-worn reference (PALbatch CREA algorithm), stratified by BMI category

(a) Performance in Normal/Underweight participants (BMI < 25 kg·m$^{-2}$)

| Model | Accuracy | F1 | Cohen's Kappa | Sensitivity | Specificity |
|---|---|---|---|---|---|
| **OxWearables - Accelerometer** | 0.849 ± 0.043 | 0.827 ± 0.055 | 0.691 ± 0.090 | 0.873 ± 0.064 | 0.830 ± 0.064 |
| **OxWearables - ActiNet** | 0.876 ± 0.042 | 0.854 ± 0.053 | 0.744 ± 0.088 | 0.883 ± 0.060 | 0.869 ± 0.055 |
| **Actimetric - Trost** | 0.735 ± 0.058 | 0.734 ± 0.063 | 0.483 ± 0.106 | 0.877 ± 0.060 | 0.633 ± 0.091 |
| **GGIR - Bakrania ENMO (32.6mg)** | 0.805 ± 0.048 | 0.788 ± 0.060 | 0.609 ± 0.098 | 0.877 ± 0.067 | 0.753 ± 0.071 |
| **GGIR - default ENMO (40mg)** | 0.801 ± 0.049 | 0.791 ± 0.058 | 0.605 ± 0.097 | 0.910 ± 0.055 | 0.721 ± 0.074 |
| **GGIR - Fraysse ENMOabs (62.5mg)** | 0.804 ± 0.048 | 0.791 ± 0.058 | 0.608 ± 0.096 | 0.895 ± 0.058 | 0.7370.073 |

Values represent mean ± standard deviation of per-participant performance.

(b) Performance in Overweight participants (BMI 25-30 kg·m$^{-2}$)

| Model | Accuracy | F1 | Cohen's Kappa | Sensitivity | Specificity |
|---|---|---|---|---|---|
| **OxWearables - Accelerometer** | 0.843 ± 0.047 | 0.824 ± 0.052 | 0.681 ± 0.089 | 0.876 ± 0.056 | 0.818 ± 0.072 |
| **OxWearables - ActiNet** | 0.869 ± 0.046 | 0.850 ± 0.052 | 0.733 ± 0.089 | 0.886 ± 0.052 | 0.857 ± 0.068 |
| **Actimetric - Trost** | 0.730 ± 0.059 | 0.733 ± 0.064 | 0.473 ± 0.107 | 0.887 ± 0.049 | 0.614 ± 0.093 |
| **GGIR - Bakrania ENMO (32.6mg)** | 0.799 ± 0.044 | 0.785 ± 0.055 | 0.597 ± 0.088 | 0.879 ± 0.057 | 0.739 ± 0.071 |
| **GGIR - default ENMO (40mg)** | 0.794 ± 0.046 | 0.787 ± 0.054 | 0.592 ± 0.088 | 0.913 ± 0.049 | 0.706 ± 0.074 |
| **GGIR - Fraysse ENMOabs (62.5mg)** | 0.797 ± 0.046 | 0.787 ± 0.055 | 0.596 ± 0.089 | 0.898 ± 0.052 | 0.723 ± 0.073 |

Values represent mean ± standard deviation of per-participant performance.

(c) Performance in Obese participants (BMI ≥ 30 $kg \cdot m^{-2}$)

| Model | Accuracy | F1 | Cohen's Kappa | Sensitivity | Specificity |
|---|---|---|---|---|---|
| **OxWearables - Accelerometer** | 0.827 ± 0.055 | 0.813 ± 0.067 | 0.652 ± 0.108 | 0.870 ± 0.059 | 0.795 ± 0.080 |
| **OxWearables - ActiNet** | 0.854 ± 0.054 | 0.840 ± 0.065 | 0.706 ± 0.108 | 0.889 ± 0.056 | 0.830 ± 0.075 |
| **Actimetric - Trost** | 0.732 ± 0.063 | 0.742 ± 0.070 | 0.477 ± 0.110 | 0.888 ± 0.056 | 0.611 ± 0.091 |
| **GGIR - Bakrania ENMO (32.6mg)** | 0.784 ± 0.058 | 0.777 ± 0.069 | 0.570 ± 0.110 | 0.872 ± 0.058 | 0.716 ± 0.083 |
| **GGIR - default ENMO (40mg)** | 0.782 ± 0.059 | 0.783 ± 0.068 | 0.570 ± 0.110 | 0.908 ± 0.049 | 0.684 ± 0.085 |
| **GGIR - Fraysse ENMOabs (62.5mg)** | 0.783 ± 0.059 | 0.781 ± 0.068 | 0.572 ± 0.110 | 0.893 ± 0.053 | 0.699 ± 0.085 |

Values represent mean ± standard deviation of per-participant performance.

**Supplementary Figure 6:** Distribution of epoch-level sedentary classification performance across six wrist-worn accelerometers relative to a thigh worn reference, stratified by BMI category

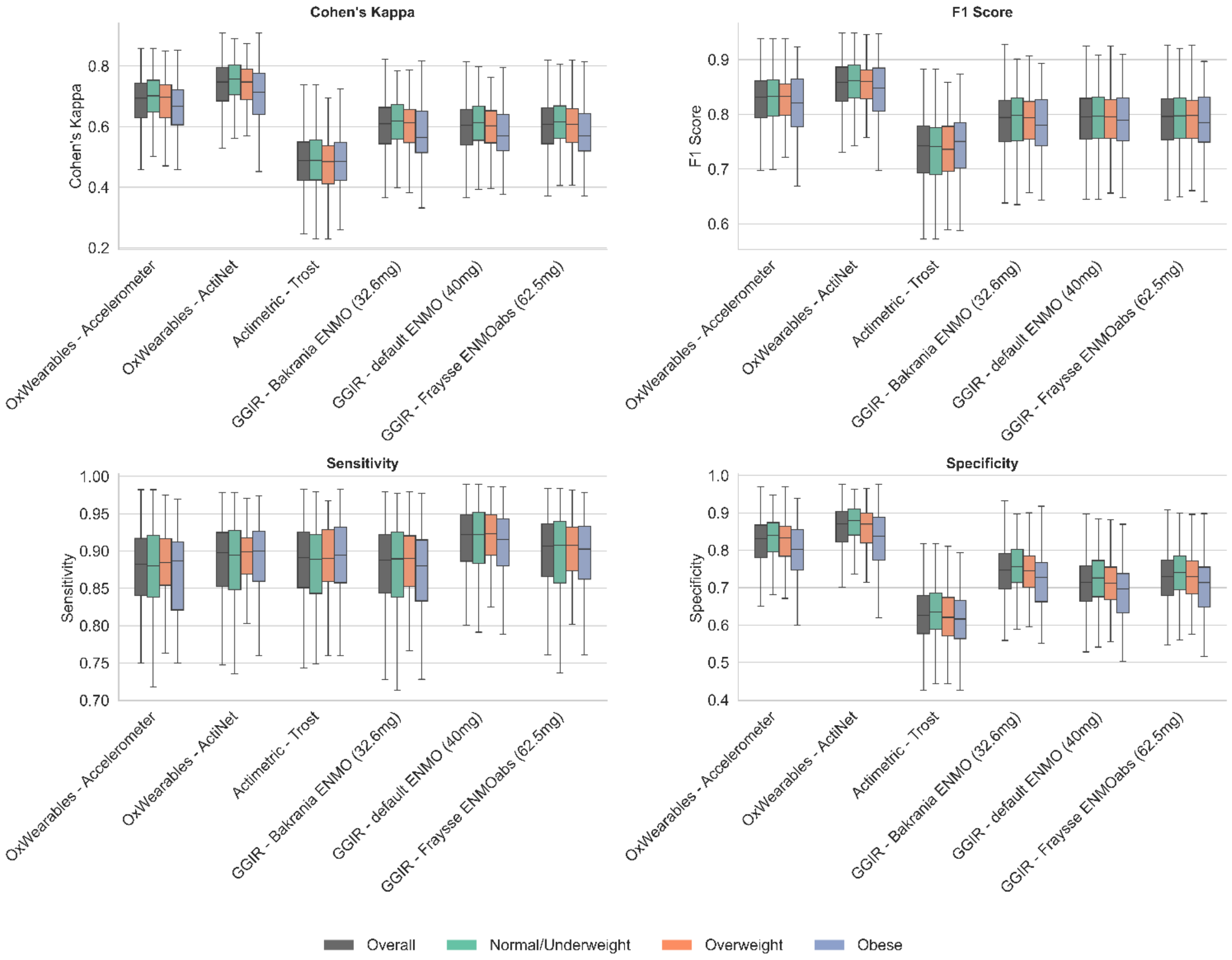

Wrist worn models compared to thigh PALbatch CREA algorithm reference

### Recruitment centre

Supplementary Table 6(a)-(b): Epoch-level sedentary classification performance of six wrist-worn accelerometer models relative to a thigh-worn reference (PALbatch CREA algorithm), stratified by recruitment centre

(a) Performance in participants recruited at Leicester

| Model | Accuracy | F1 | Cohen's Kappa | Sensitivity | Specificity |
|---|---|---|---|---|---|
| **OxWearables - Accelerometer** | 0.843 ± 0.047 | 0.824 ± 0.057 | 0.681 ± 0.095 | 0.875 ± 0.060 | 0.818 ± 0.070 |
| **OxWearables - ActiNet** | 0.869 ± 0.047 | 0.851 ± 0.055 | 0.732 ± 0.096 | 0.886 ± 0.056 | 0.856 ± 0.066 |
| **Actimetric - Trost** | 0.737 ± 0.056 | 0.740 ± 0.065 | 0.487 ± 0.102 | 0.886 ± 0.053 | 0.627 ± 0.084 |
| **GGIR - Bakrania ENMO (32.6mg)** | 0.799 ± 0.050 | 0.786 ± 0.060 | 0.597 ± 0.099 | 0.878 ± 0.059 | 0.739 ± 0.076 |
| **GGIR - default ENMO (40mg)** | 0.795 ± 0.051 | 0.789 ± 0.059 | 0.593 ± 0.100 | 0.913 ± 0.049 | 0.706 ± 0.078 |
| **GGIR - Fraysse ENMOabs (62.5mg)** | 0.797 ± 0.051 | 0.788 ± 0.059 | 0.595 ± 0.100 | 0.897 ± 0.054 | 0.7210.078 |

Values represent mean ± standard deviation of per-participant performance.

(b) Performance in participants recruited at Salford

| Model | Accuracy | F1 | Cohen's Kappa | Sensitivity | Specificity |
|---|---|---|---|---|---|
| **OxWearables - Accelerometer** | 0.842 ± 0.049 | 0.821 ± 0.058 | 0.678 ± 0.094 | 0.871 ± 0.062 | 0.821 ± 0.073 |
| **OxWearables - ActiNet** | 0.869 ± 0.046 | 0.849 ± 0.056 | 0.733 ± 0.091 | 0.883 ± 0.058 | 0.860 ± 0.065 |
| **Actimetric - Trost** | 0.726 ± 0.064 | 0.728 ± 0.065 | 0.466 ± 0.113 | 0.876 ± 0.061 | 0.617 ± 0.103 |
| **GGIR - Bakrania ENMO (32.6mg)** | 0.799 ± 0.050 | 0.784 ± 0.062 | 0.598 ± 0.098 | 0.873 ± 0.067 | 0.745 ± 0.074 |
| **GGIR - default ENMO (40mg)** | 0.795 ± 0.051 | 0.787 ± 0.059 | 0.595 ± 0.096 | 0.907 ± 0.056 | 0.714 ± 0.077 |
| **GGIR - Fraysse ENMOabs (62.5mg)** | 0.798 ± 0.050 | 0.787 ± 0.059 | 0.599 ± 0.095 | 0.893 ± 0.057 | 0.730 ± 0.076 |

Values represent mean ± standard deviation of per-participant performance.

**Supplementary Figure 7:** Distribution of epoch-level sedentary classification performance across six wrist-worn accelerometers relative to a thigh worn reference, stratified by recruitment centre

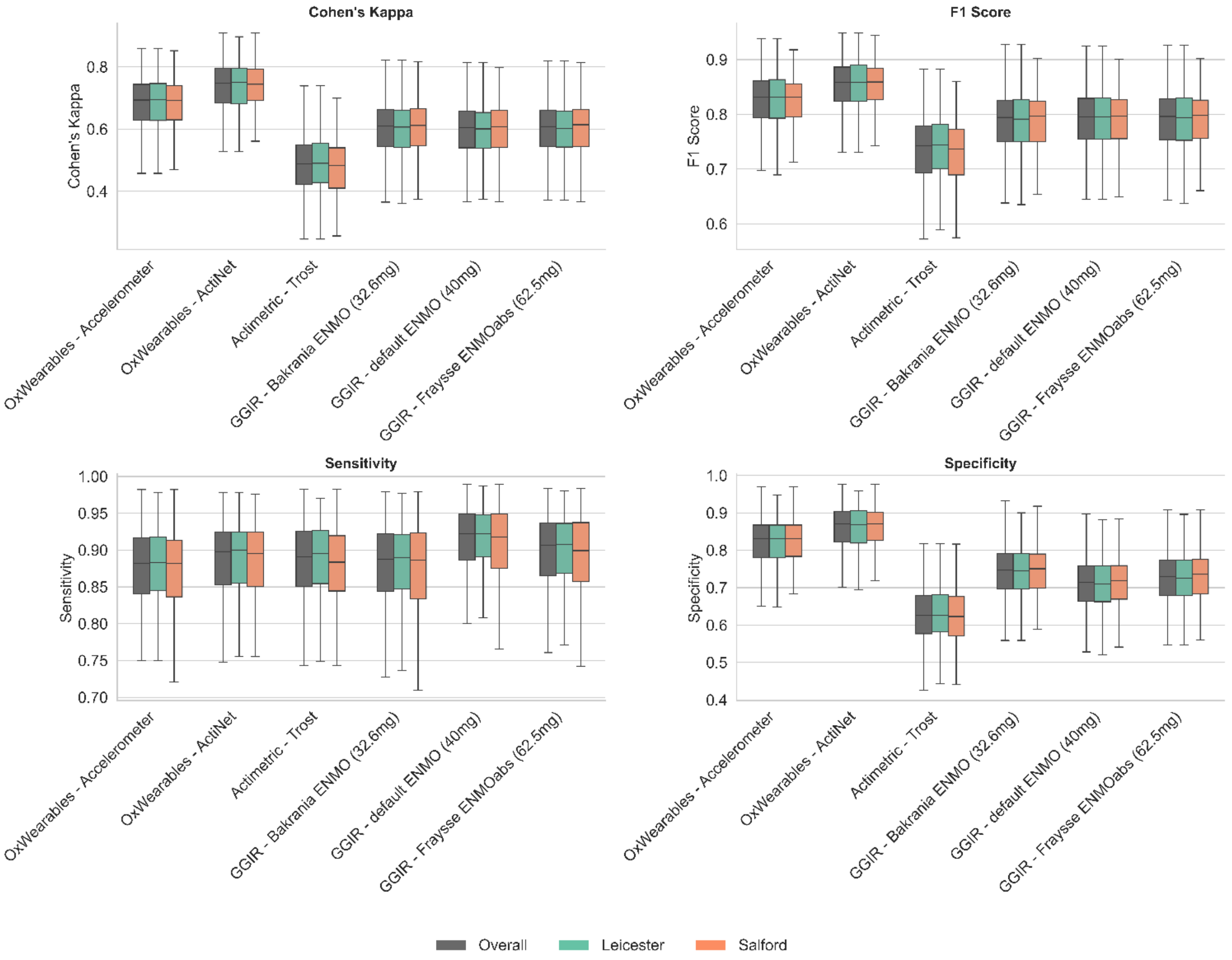

Wrist worn models compared to thigh PALbatch CREA algorithm reference

### Season of accelerometer wear

Supplementary Table 7(a)-(d): Epoch-level sedentary classification performance of six wrist-worn accelerometer models relative to a thigh-worn reference (PALbatch CREA algorithm), stratified by season of accelerometer wear.

(a) Performance in participants monitored during Spring (March -May)

| Model | Accuracy | F1 | Cohen's Kappa | Sensitivity | Specificity |
|---|---|---|---|---|---|
| **OxWearables - Accelerometer** | 0.838 ± 0.044 | 0.814 ± 0.050 | 0.670 ± 0.085 | 0.865 ± 0.062 | 0.819 ± 0.064 |
| **OxWearables - ActiNet** | 0.861 ± 0.048 | 0.837 ± 0.054 | 0.714 ± 0.095 | 0.873 ± 0.065 | 0.852 ± 0.066 |
| **Actimetric - Trost** | 0.729 ± 0.055 | 0.726 ± 0.063 | 0.472 ± 0.100 | 0.879 ± 0.057 | 0.624 ± 0.087 |
| **GGIR - Bakrania ENMO (32.6mg)** | 0.790 ± 0.055 | 0.772 ± 0.058 | 0.580 ± 0.106 | 0.868 ± 0.058 | 0.735 ± 0.085 |
| **GGIR - default ENMO (40mg)** | 0.785 ± 0.056 | 0.776 ± 0.057 | 0.575 ± 0.105 | 0.903 ± 0.051 | 0.702 ± 0.089 |
| **GGIR - Fraysse ENMOabs (62.5mg)** | 0.788 ± 0.056 | 0.775 ± 0.057 | 0.579 ± 0.106 | 0.887 ± 0.054 | 0.7180.088 |

Values represent mean ± standard deviation of per-participant performance.

(b) Performance in participants monitored during Summer (June-August)

| Model | Accuracy | F1 | Cohen's Kappa | Sensitivity | Specificity |
|---|---|---|---|---|---|
| **OxWearables - Accelerometer** | 0.842 ± 0.053 | 0.823 ± 0.060 | 0.678 ± 0.104 | 0.873 ± 0.060 | 0.817 ± 0.079 |
| **OxWearables - ActiNet** | 0.869 ± 0.051 | 0.851 ± 0.058 | 0.732 ± 0.101 | 0.885 ± 0.056 | 0.857 ± 0.072 |
| **Actimetric - Trost** | 0.732 ± 0.068 | 0.736 ± 0.068 | 0.478 ± 0.121 | 0.886 ± 0.056 | 0.619 ± 0.108 |
| **GGIR - Bakrania ENMO (32.6mg)** | 0.800 ± 0.050 | 0.786 ± 0.061 | 0.599 ± 0.101 | 0.878 ± 0.062 | 0.741 ± 0.077 |
| **GGIR - default ENMO (40mg)** | 0.795 ± 0.052 | 0.789 ± 0.060 | 0.593 ± 0.102 | 0.912 ± 0.052 | 0.707 ± 0.080 |
| **GGIR - Fraysse ENMOabs (62.5mg)** | 0.798 ± 0.051 | 0.788 ± 0.060 | 0.597 ± 0.101 | 0.897 ± 0.055 | 0.723 ± 0.080 |

Values represent mean ± standard deviation of per-participant performance.

(c) Performance in participants monitored during Autumn (September-November)

| Model | Accuracy | F1 | Cohen's Kappa | Sensitivity | Specificity |
|---|---|---|---|---|---|
| **OxWearables - Accelerometer** | 0.843 ± 0.047 | 0.823 ± 0.060 | 0.682 ± 0.094 | 0.872 ± 0.064 | 0.823 ± 0.067 |
| **OxWearables - ActiNet** | 0.871 ± 0.044 | 0.851 ± 0.057 | 0.735 ± 0.090 | 0.885 ± 0.061 | 0.861 ± 0.060 |
| **Actimetric - Trost** | 0.734 ± 0.054 | 0.734 ± 0.064 | 0.479 ± 0.097 | 0.875 ± 0.062 | 0.630 ± 0.078 |
| **GGIR - Bakrania ENMO (32.6mg)** | 0.800 ± 0.054 | 0.785 ± 0.067 | 0.600 ± 0.105 | 0.875 ± 0.070 | 0.745 ± 0.074 |
| **GGIR - default ENMO (40mg)** | 0.796 ± 0.054 | 0.789 ± 0.064 | 0.597 ± 0.102 | 0.908 ± 0.059 | 0.715 ± 0.077 |
| **GGIR - Fraysse ENMOabs (62.5mg)** | 0.799 ± 0.053 | 0.789 ± 0.064 | 0.601 ± 0.101 | 0.894 ± 0.058 | 0.731 ± 0.075 |

Values represent mean ± standard deviation of per-participant performance.

(d) Performance in participants monitored during Winter (December-February)

| Model | Accuracy | F1 | Cohen's Kappa | Sensitivity | Specificity |
|---|---|---|---|---|---|
| **OxWearables - Accelerometer** | 0.845 ± 0.039 | 0.827 ± 0.049 | 0.684 ± 0.078 | 0.879 ± 0.057 | 0.817 ± 0.060 |
| **OxWearables - ActiNet** | 0.871 ± 0.039 | 0.855 ± 0.048 | 0.737 ± 0.079 | 0.891 ± 0.050 | 0.855 ± 0.058 |
| **Actimetric - Trost** | 0.736 ± 0.050 | 0.740 ± 0.059 | 0.483 ± 0.090 | 0.887 ± 0.047 | 0.621 ± 0.075 |
| **GGIR - Bakrania ENMO (32.6mg)** | 0.800 ± 0.041 | 0.788 ± 0.051 | 0.599 ± 0.080 | 0.879 ± 0.052 | 0.739 ± 0.067 |
| **GGIR - default ENMO (40mg)** | 0.798 ± 0.042 | 0.794 ± 0.051 | 0.600 ± 0.081 | 0.915 ± 0.044 | 0.709 ± 0.068 |
| **GGIR - Fraysse ENMOabs (62.5mg)** | 0.798 ± 0.042 | 0.791 ± 0.052 | 0.599 ± 0.082 | 0.898 ± 0.050 | 0.722 ± 0.067 |

Values represent mean ± standard deviation of per-participant performance.

**Supplementary Figure 8:** Distribution of epoch-level sedentary classification performance across six wrist-worn accelerometers relative to a thigh worn reference, stratified by wear season

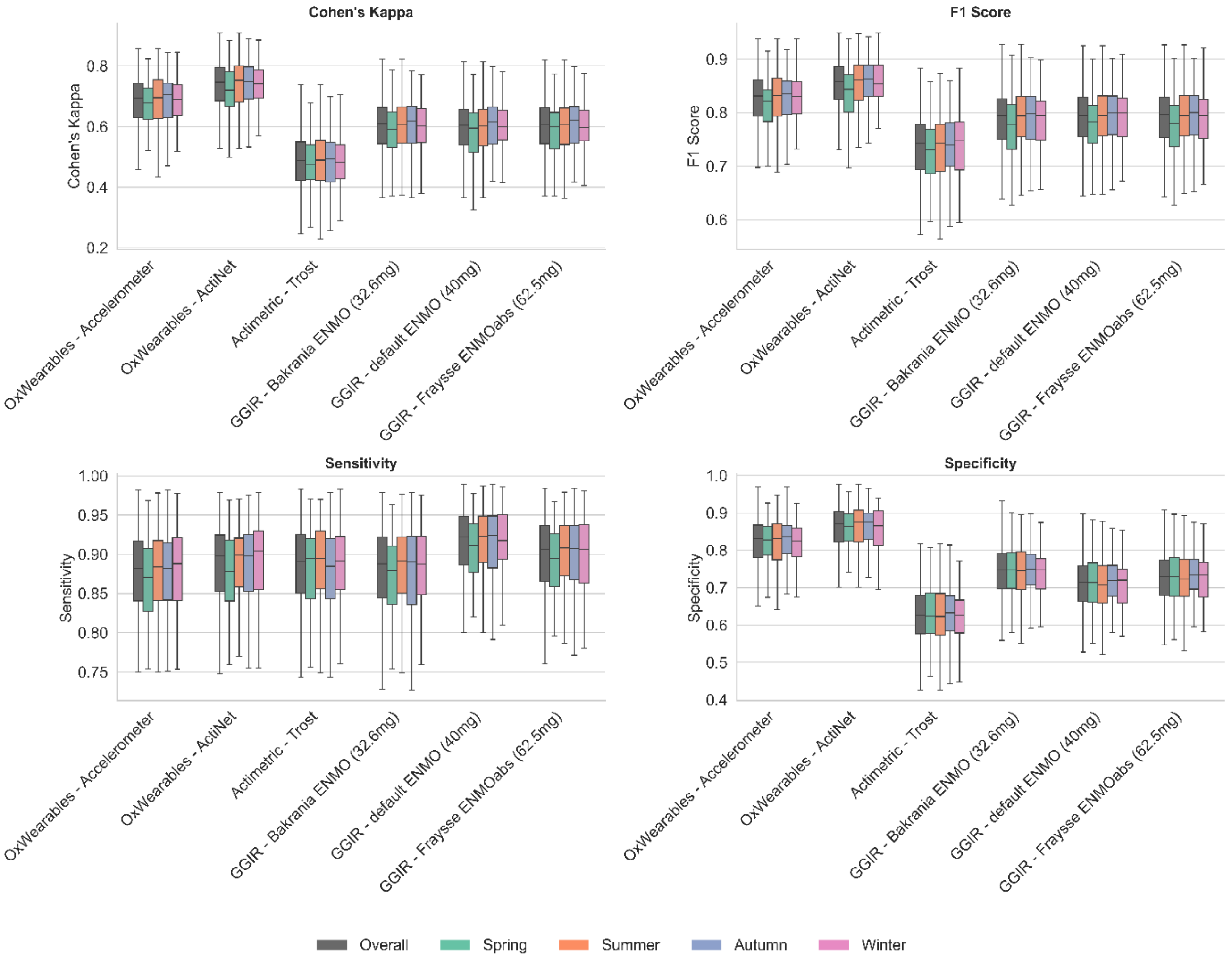

Wrist worn models compared to thigh PALbatch CREA algorithm reference.
Spring: March-May; Summer: June-August; Autumn: September-November; Winter: December-February

### Sensitivity Analysis

Supplementary Table 8: Epoch-level sedentary classification performance of six wrist-worn accelerometer models relative to a thigh-worn reference (PALbatch CREA algorithm), restricted to waking hours (08:00-22:00)

| Model | Accuracy | F1 | Cohen's Kappa | Sensitivity | Specificity |
|---|---|---|---|---|---|
| **OxWearables - Accelerometer** | 0.815 ± 0.057 | 0.854 ± 0.054 | 0.584 ± 0.117 | 0.889 ± 0.057 | 0.684 ± 0.119 |

| | | | | | |
|---|---|---|---|---|---|
| **OxWearables - ActiNet** | **0.853 ± 0.055** | **0.883 ± 0.051** | **0.675 ± 0.116** | 0.901 ± 0.052 | **0.769 ± 0.112** |
| **Actimetric - Trost** | 0.696 ± 0.070 | 0.783 ± 0.063 | 0.280 ± 0.119 | 0.891 ± 0.055 | 0.368 ± 0.119 |
| **GGIR - Bakrania ENMO (32.6mg)** | 0.766 ± 0.063 | 0.823 ± 0.059 | 0.465 ± 0.117 | 0.888 ± 0.060 | 0.559 ± 0.114 |
| **GGIR - default ENMO (40mg)** | 0.762 ± 0.065 | 0.826 ± 0.058 | 0.442 ± 0.117 | **0.922 ± 0.048** | 0.490 ± 0.116 |
| **GGIR - Fraysse ENMOabs (62.5mg)** | 0.765 ± 0.064 | 0.825 ± 0.058 | 0.455 ± 0.117 | 0.907 ± 0.051 | 0.523 ± 0.117 |

Performance scores are reported as mean ± standard deviation across all participants in the study. High score model for respective performance metric is indicated in bold.

**Supplementary Figure 9:** Distribution of epoch-level sedentary classification performance across six wrist-worn accelerometer models relative to a thigh-worn reference (PALbatch CREA algorithm), restricted to waking hours (08:00–22:00)

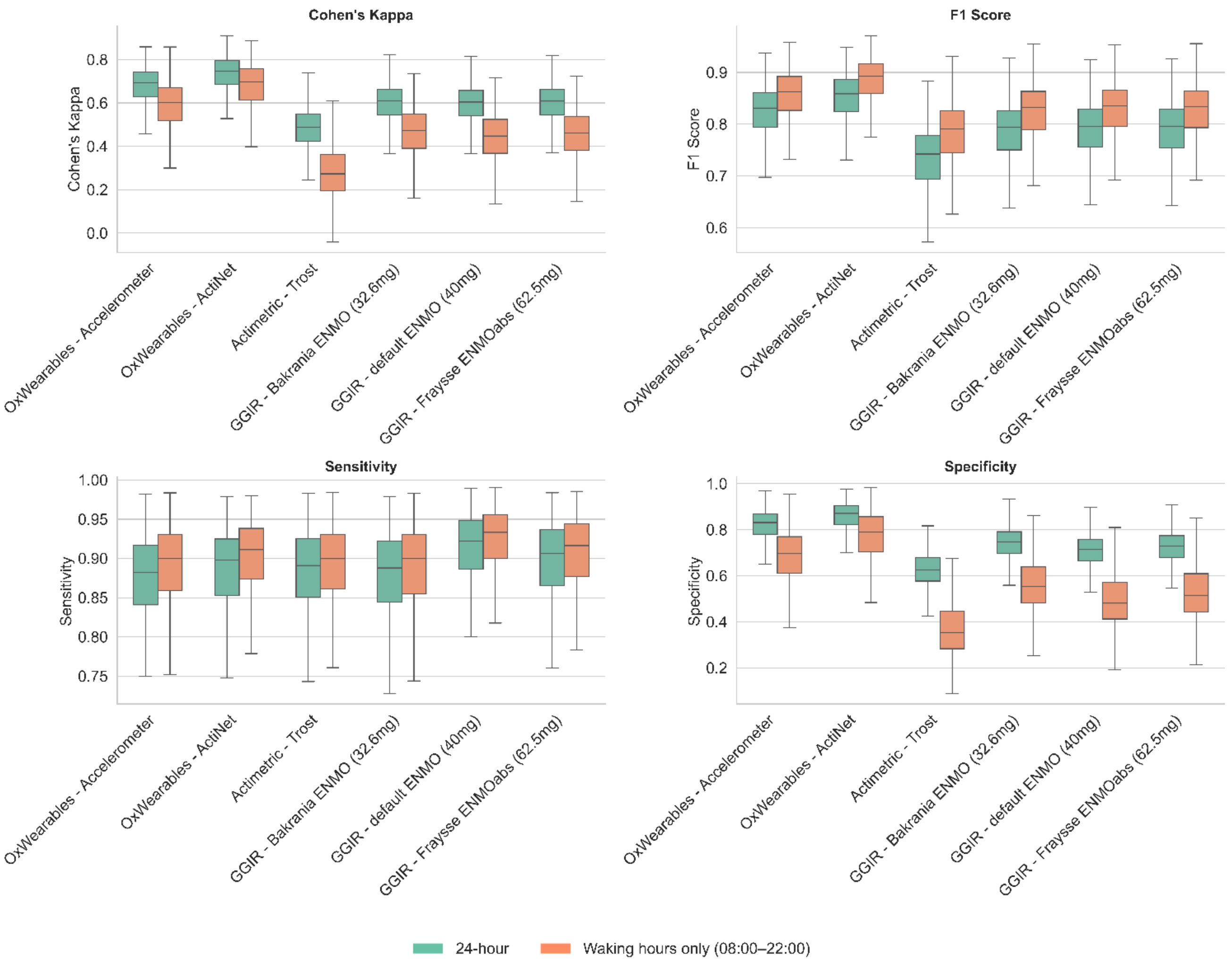

Supplementary Table 9(a)-(c): Agreement in wrist-derived sedentary metrics across six wrist-worn accelerometer models relative to a thigh-worn reference (PALbatch CREA algorithm), restricted to waking hours (08:00-22:00).

(a) Average daily sedentary time (hours/day)

| | Mean (SD) | RMSE | MAE | Pearson's r |
|---|---|---|---|---|
| **PALBatch - CREA [ref]** | 8.354 (1.287) | - | - | - |
| **OxWearables - Accelerometer** | 9.004 (1.374) | 1.143 | 0.852 | 0.752 |
| **OxWearables - ActiNet** | 8.676 (1.334) | 0.859 | 0.596 | 0.816 |
| **Actimetric - Trost** | 10.583 (1.279) | 2.529 | 2.250 | 0.566 |
| **GGIR - Bakrania ENMO (32.6mg)** | 9.619 (1.310) | 1.619 | 1.342 | 0.697 |
| **GGIR - default ENMO (40mg)** | 10.245 (1.282) | 2.143 | 1.904 | 0.691 |
| **GGIR - Fraysse ENMOabs (62.5mg)** | 9.955 (1.288) | 1.893 | 1.628 | 0.691 |

Prolonged sedentary bouts are defined as detected periods of sedentary behaviour, lasting at least 30 minutes, with no gap in sedentary behaviour for over 90 seconds, and minimum proportion of detected sedentary behaviour of 80%. RMSE – root mean squared error,
MAE – mean absolute error, Pearson's r – Pearson's correlation coefficient, SD – standard deviation.

(b) Average daily number of prolonged sedentary bouts (count/day)

| | Mean (SD) | RMSE | MAE | Pearson's r |
|---|---|---|---|---|
| **PALBatch - CREA [ref]** | 5.66 (1.04) | - | - | - |
| **OxWearables - Accelerometer** | 5.41 (0.90) | 1.01 | 0.78 | 0.500 |
| **OxWearables - ActiNet** | 5.64 (0.97) | 0.78 | 0.59 | 0.698 |
| **Actimetric - Trost** | 4.89 (0.94) | 1.50 | 1.18 | 0.143 |
| **GGIR - Bakrania ENMO (32.6mg)** | 5.57 (0.91) | 1.00 | 0.78 | 0.484 |
| **GGIR - default ENMO (40mg)** | 5.56 (0.95) | 1.14 | 0.87 | 0.354 |
| **GGIR - Fraysse ENMOabs (62.5mg)** | 5.59 (0.90) | 1.03 | 0.80 | 0.442 |

Prolonged sedentary bouts are defined as detected periods of sedentary behaviour, lasting at least 30 minutes, with no gap in sedentary behaviour for over 90 seconds, and minimum proportion of detected sedentary

behaviour of 80%. RMSE – root mean squared error,
MAE – mean absolute error, Pearson's r – Pearson's correlation coefficient, SD – standard deviation.

(c) Proportion of sedentary time accumulated in prolonged sedentary bouts (%)

| | Mean (SD) | RMSE | MAE | Pearson's r |
|---|---|---|---|---|
| **PALBatch - CREA [ref]** | 73.0 (9.3) | - | - | - |
| **OxWearables - Accelerometer** | 84.2 (6.3) | 13.3 | 11.3 | 0.625 |
| **OxWearables - ActiNet** | 80.0 (7.2) | 9.3 | 7.4 | 0.752 |
| **Actimetric - Trost** | 91.8 (10.2) | 22.1 | 19.4 | 0.273 |
| **GGIR - Bakrania ENMO (32.6mg)** | 80.1 (9.2) | 11.2 | 8.8 | 0.552 |
| **GGIR - default ENMO (40mg)** | 84.5 (8.2) | 14.5 | 12.0 | 0.503 |
| **GGIR - Fraysse ENMOabs (62.5mg)** | 82.9 (8.5) | 13.1 | 10.6 | 0.535 |

Prolonged sedentary bouts are defined as detected periods of sedentary behaviour, lasting at least 30 minutes, with no gap in sedentary behaviour for over 90 seconds, and minimum proportion of detected sedentary behaviour of 80%. RMSE – root mean squared error,
MAE – mean absolute error, Pearson's r – Pearson's correlation coefficient, SD – standard deviation.
